# Evaluation of symptom checker formats to support health literacy and trust in AI: Results from an online randomised-controlled trial

**DOI:** 10.64898/2026.03.11.26347036

**Authors:** Julie Ayre, Karen Gallagher, Jenna Smith, Kirsten Ingwersen, Claire Hudson, Angelica Scott, Alison Woods, Christopher Ng, Yuanee Wickramasinghe, Ivan CK Ma, Waren Nadesan, Gauri Kapoor, Geoffrey Edlund, Lorna Butters, Trang Vu, Kirsten J McCaffery

## Abstract

**Objectives:** Evaluate the impact of online symptom checker formats on symptom management knowledge, symptom checker trust and acceptability, and behavioural intentions.

**Design:** Two 5-arm parallel-group online randomised controlled trials.

**Setting:** Online survey.

**Participants:** Sample of 2062 Australian adults recruited through online research panel in June 2025. Almost half identified as man/male (49%) and woman/female (51%), median age 49 years (IQR=28). Participants viewed a hypothetical health scenario (fever and vomiting) followed by a screenshot of an online symptom checker from national health service provider, healthdirect.

**Interventions:** Participants were randomised to symptom acuity level (low: self-care at home; or moderate: see a General Practitioner (GP) in 24 hours) and one of five symptom checker formats. The standard format showed the existing healthdirect symptom checker advice. The remaining formats were ‘AI-enhanced’ versions. These included an AI-enhanced version with e.g. more tailored advice, rationale for acuity level, and AI disclosure statement. The other AI-enhanced formats had additional features: numbered steps, multimedia, and more detailed information about the use of AI.

**Main outcome measures:** Primary outcomes were intentions to follow the symptom checker’s self-care advice and intentions to see a GP in 24 hours. Secondary outcomes were trust in advice, knowledge of symptom management, and acceptability of the tool. All outcomes were assessed immediately post-intervention; knowledge was also assessed after 2 weeks.

**Results:** When advised to self-care at home, the AI-enhanced groups reported lower intentions to see a GP in 24 hours (median 3.00 out of 5), compared to the standard (original) tool (median 4.00; adjusted p = 0.003). There were no other significant effects on intentions. Immediately following the intervention, participants who viewed an AI-enhanced format reported greater knowledge about how to manage current and changing symptoms, across both acuity levels (adjusted p’s <0.001). Knowledge gains were not sustained at 2 week follow-up. There were no significant effects on trust or acceptability.

**Conclusions:** Participants who viewed the more tailored information in the AI-enhanced formats demonstrated stronger knowledge for managing symptoms than those who viewed the standard format. There was also some evidence that an AI-enhanced format may be more effective at reducing use of primary care for symptoms that can be managed at home. Trust and acceptability were high across formats, and the explicit use of AI did not impact significantly on these outcomes. Future research should investigate these formats using interactive prototypes across a wider variety of health contexts.

**Registration:** ACTRN ACTRN12625000474459p

**Key messages:** *What is already known on this topic:* Although online, evidence-based symptom checkers have been widely available from reputable health organisations for over a decade, they often face poor uptake and may not adequately meet health literacy needs of diverse users.

*What this study adds:* Symptom checker features that could be implemented with AI, such as tailored information and a clear rationale for triage advice, may help support appropriate symptom management. Statements about the tool’s use of AI did not appear to impact trust or acceptability of the symptom checker tool.

*How this study might affect research, practice or policy:* Findings from this study suggest that using AI to enhance symptom checker advice may not impact negatively on trust and acceptability of the tool, and may improve appropriate symptom management. Further research is needed to investigate AI-enhanced symptom checker formats using interactive prototypes across a wider variety of health contexts.

## INTRODUCTION

Online symptom checkers are evidence-informed tools that can help people manage their symptoms and seek appropriate care. These tools have been publicly available for over a decade, via mainstream services such as the UK’s National Health Service and Australia’s healthdirect.^1^ They are increasingly relevant in the current policy climate which emphasises reducing unnecessary and low-value care, and providing accessible, trustworthy health advice.^2^ ^3^ Though several studies have highlighted potential issues with accuracy, or argue their advice is too risk-averse to substantially reduce health system burden,^4^ ^5^ online symptom checkers remain an important scalable tool for timely tailored health advice, particularly in the face of general-purpose artificial intelligence (AI) tools such as ChatGPT.^6–9^

Research shows that people are generally supportive of online symptom checkers, ^5^ ^10–12^ but there is also mounting evidence that they could better meet community needs. For example, even if people use an online symptom checker, they may not follow the triage advice. Whilst some studies suggest that intentions to follow symptom checker advice are high (e.g. 87% of 634 UK adults reported following the advice most/all of the time)^11^, translation into action is likely much lower.^13^ ^14^ Following advice may also be lower when advice is more urgent, although evidence for this is mixed.^13–15^

Current designs for online symptom checkers may also inadvertently contribute to health inequity and the digital divide. For example, there is growing evidence that users are more likely to be female, educated, white, and to have stronger digital literacy skills.^4^ ^5^ ^16^ People who find these tools easier to use are up to 8 times more likely to use them, ^11^ with qualitative research suggesting that usability may be hindered by poor implementation of health literacy principles, particularly around the use of medical jargon.^5 10 17 18^

As is the case with other health information sources, trust likely plays a key role in use of online symptom checkers.^19^ Some barriers to trust are common to most digital health tools e.g. lack of endorsement by trusted authorities and concerns about data privacy.^5^ Other barriers to trust are more specific, such as users feeling that their symptoms were poorly represented by the tool or that the health advice was not plausible, vague, or lacked transparency about the tool’s decision-making process.^5^ ^10^ ^18^ ^20^ ^21^

Recent technological advances may provide an opportunity to address some of these issues and develop online symptom checkers that better meet the needs of diverse communities. For example, tools that integrate generative AI may be able to more flexibly tailor content for users,^22^ and there is growing evidence that generative AI may be an effective tool for creating health materials that more closely adhere to plain language guidelines.^23^ ^24^ These potential benefits must be weighed against potential risks to safety and erosion of trust, particularly if the symptom checker is delivered through a reputable organisation.^25^ ^26^

Although there is a well-established literature evaluating the accuracy of AI triage tools,^4^ ^5^ only a few studies have investigated the use of AI to make symptom checker communication more accessible and improve end-user experience. Some limited evidence suggests people are just as trusting of symptom checker tools that are explicit about their use of AI as those that are not.^27^ There is also some evidence that the manner in which a symptom checker explains triage advice can impact on trust, with one study reporting greater trust in advice for managing unfamiliar symptoms when there was a clear link between symptom input and advice, compared to when no explanation was provided.^28^ However, both studies were conducted prior to the release of publicly available generative AI tools such as ChatGPT, and it is likely that the public’s relationship with and expectations of AI have changed substantially.

To address these research gaps, this study aimed to evaluate the impacts of different symptom checker formats relating to health literacy and potential AI-enhancements on symptom management knowledge, symptom checker trust and acceptability, and behavioural intentions. This research question was assessed across low and moderate symptom acuity levels i.e. situations where symptoms can be managed through self-care (low acuity) or by accessing primary care in 24 hours (moderate acuity).

## METHODS

### Study Design

This study involved two 5-arm parallel-group online randomised controlled trials (Figure 1). Participants were randomly allocated to an intervention group using survey platform Qualtrics’ randomizer function, a pseudorandom number generator based on the Mersenne Twister. The trial was prospectively registered with the Australian New Zealand Clinical Trials Registry (ACTRN12625000474459p) in May 2024, and approved by the University of Sydney Research Ethics Committee (2025/HE000143).

**Figure 1.**
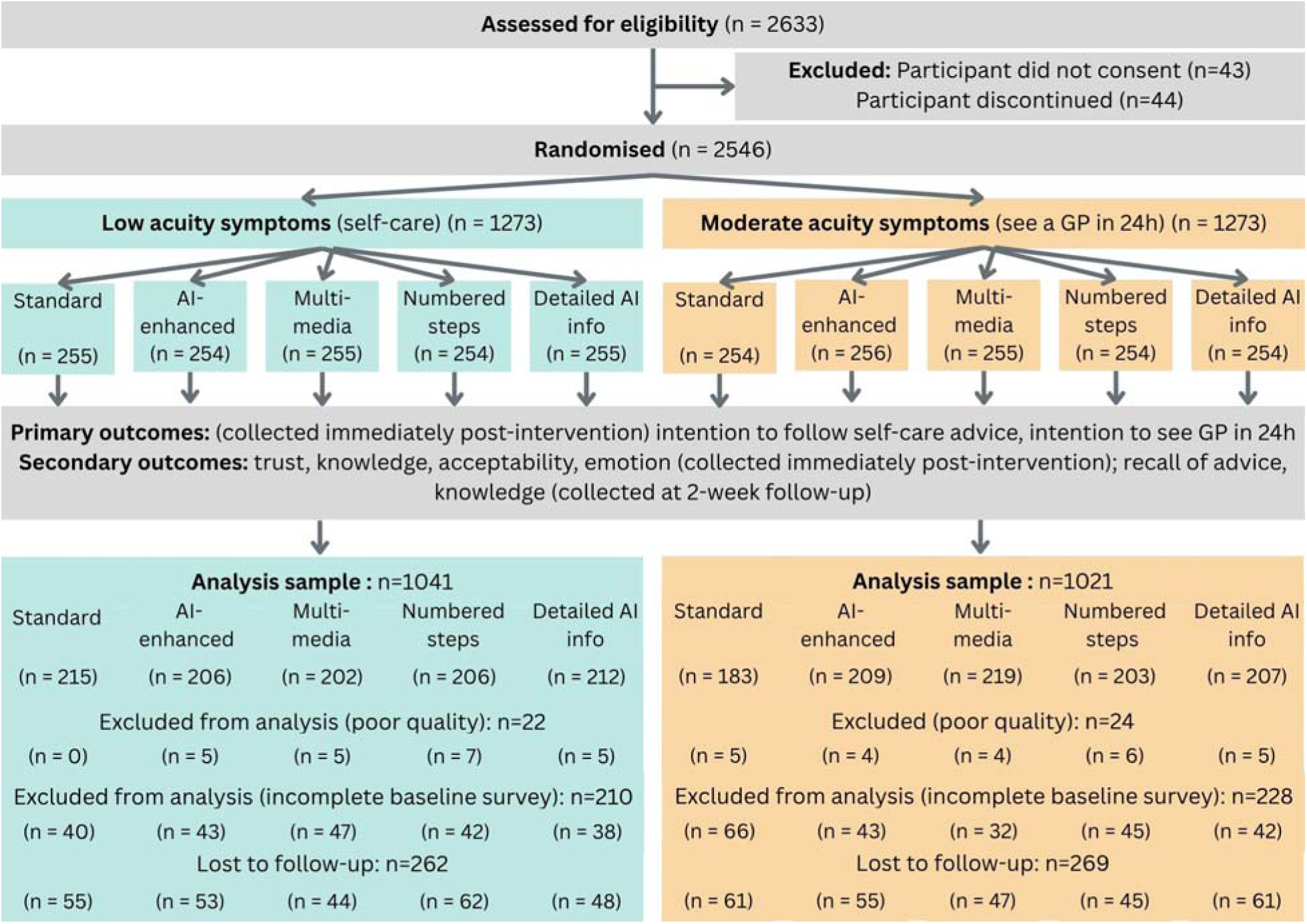
Study flow diagram. *Note*. Participants were excluded from the analysis if they were considered ‘speeders’ i.e. completed the survey in less than 5 minutes.

### Digital health context

Healthdirect Australia is a national government-funded health service that provides free health advice and information via a website, app and telephone helpline. The healthdirect online symptom checker was launched in 2014 and is based on an established evidence-based clinical decision support technology. At the time of writing, the existing symptom checker did not use generative AI. Advice is tailored according to triage category (e.g. self-care, see a general practitioner (GP) in 24 hours, go to emergency) rather than specific symptoms. The tool can also connect users with local service information and availability.

Healthdirect also hosts more than 3000 web pages on health topics and medicines information. This provides a curated knowledge library that an AI model could draw from, allowing the symptom checker to provide more tailored advice. For example, retrieval-augmented generation (RAG) frameworks can connect an AI model with external knowledge.^29^ Although using a RAG framework cannot completely ensure clinical accuracy, studies demonstrate it can considerably improve the accuracy of generative AI tools compared to those that do not provide a knowledge base.^30^ ^31^

### Participants

Eligible participants were adults (18+ years) who resided in Australia and who self-reported good English proficiency. Given that online symptom checkers may be underutilised by people who have not had opportunity for tertiary education,^5^ we used quotas to ensure that no more than 50% of the sample had a university education. Participants were recruited between June 19^th^ and 22^nd^, 2025, via an online research panel comprising more than 550,000 Australian adults. Participants were aware that they would be allocated to different symptom checker formats, but not that some formats incorporated generative AI. Participants completed the follow-up survey at least two weeks later.

### Procedure

The survey was hosted using Qualtrics software. After consenting, participants completed questions about demographics, health literacy,^32^ and digital health literacy.^33^ The survey items also asked about awareness and previous use of healthdirect and AI tools (Appendix B).

Participants across all intervention groups were then asked to imagine the following health scenario:

> *You currently have a fever and have vomited twice. You’re worried that you are dehydrated so you try sipping on some apple juice, which you immediately throw up again. Yesterday you had a little less energy than usual, but you were still able to complete the things you needed to do*.
>
> *You are not quite sure what to do, so you use the healthdirect symptom checker. The symptom checker is an online tool to help people decide whether to see a doctor or look after themselves at home. To help you find a local service it will also ask for your postcode, you put in Barangaroo 2000*.
>
> ***The symptom checker does not give a diagnosis and is not a substitute for professional healthcare.***

Participants were then randomised to one of 10 intervention groups (Figure 1), and were asked to view the corresponding health advice for at least 2 minutes. Immediately after viewing the advice, participants completed an attention check that asked them to identify the key symptoms described in the health scenario. Participants then completed primary and secondary outcomes.

After two weeks participants completed a second survey. This reminded them of the original health scenario, and participants were asked to recall their triage advice, and completed the knowledge outcome. At the end of the study all participants were debriefed, including a statement that the scenario was hypothetical and that they did not receive information or advice regarding their personal health situation.

### Intervention groups

All participants viewed a screenshot of the advice page (key components shown in Figure 2). Links and video content shown in the screenshot were also available to participants.

**Figure 2.**
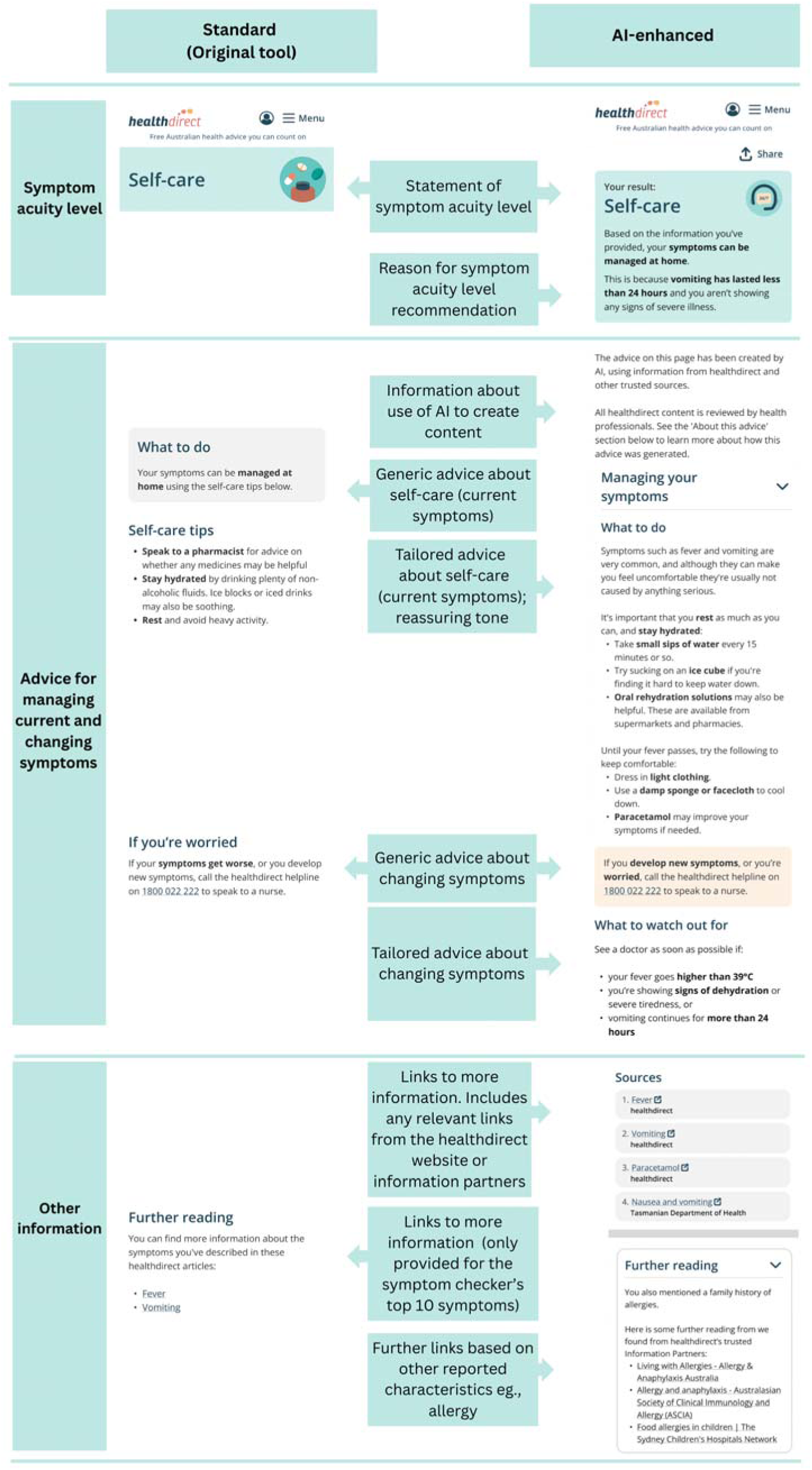
Comparison of standard and AI-enhanced groups, low symptom acuity level (self-care)

#### Symptom acuity level

##### Low acuity symptoms

Participants were advised to manage their symptoms at home (Figure 2).

##### Moderate acuity symptoms

Participants were advised to see a GP within 24 hours and provided with links to local GPs and Urgent Care Services for postcode 2000, the postcode used in the scenario (eFigure 1).

#### Symptom checker formats

##### Standard

The standard format reflected the original symptom checker tool’s content and format. Advice about symptom management was not specific to the symptoms (eFigures 2 and 3).

##### AI-enhanced

This format reflected several AI-enhancements, including advice that was tailored to specific symptoms, links to relevant information sources, a statement that the content had been generated by AI, and a reassuring tone for low acuity presentations (eFigures 4 and 5). The advice also explicitly described how the symptoms related to the acuity level. These features appeared in all groups other than the Standard group.

##### AI-enhanced + numbered steps

This format included numbered steps for following self-management advice (eFigures 6 and 7).

##### AI-enhanced + multimedia

This format incorporated a video about managing fever and an image showing the symptoms of fever (eFigures 8 and 9).

##### AI-enhanced + detailed information about AI

This format provided more detailed information about how AI was integrated into the tool (eFigures 10 and 11).

### Outcomes

All outcomes were assessed immediately following the intervention. Knowledge was also assessed at 2-week follow-up.

#### Intention to follow symptom checker advice

The primary outcome was intention to follow symptom checker advice. This was assessed using two items. The first item assessed intentions to follow the self-care advice, using a 4-point scale from *“I would follow all the advice”* to *“I would follow none of the advice”.* The second assessed intentions to see a GP in 24 hours; participants responded using a 5-point scale from “*definitely would*” to “*definitely would not*”.

#### Trust

Trust was assessed using three instruments. The first was a single-item 10-point Likert scale that asked participants to rate how trustworthy they found the advice, adapted from Mac and colleagues.^34^ Cognitive trust (4 items) and emotional trust (3 items) were each assessed using a validated instrument^35^ that has previously been used to evaluate AI-enhanced symptom checker advice.^36^ Reponses were recorded on a 7-point Likert scale.

#### Knowledge

Knowledge was assessed using purpose-built multiple-choice questions that were based on the content available in the intervention (Appendix B eTable 1). This produced two scores: one for managing current symptoms and the second for managing changing symptoms. Scores are presented as the proportion (%) of correct responses. At 2-week follow-up participants were also asked to recall the acuity level (i.e. to self-care at home for low acuity and to see a GP in 24 hours for moderate acuity).

#### Acceptability

Acceptability of the symptom checker was assessed using a 5-point Likert scale from (1) “strongly disagree” to (5) “strongly agree.” Three purpose-built statements asked about the extent that they would use the tool in the future, recommend the tool to a friend; and the extent that the tool met their expectations.

Items relating to emotional responses^37^ were also collected. They are not analysed here.

### Analysis

An intention to treat analysis was performed, comparing the 5 format groups for each symptom acuity level. Descriptive statistics are presented as medians, interquartile ranges and means for continuous variables and relative percentages for categorical variables. Statistical significance of intervention group differences was tested using Kruskal-Wallis tests comparing mean ranks, and one-way ANOVA tests where appropriate, where significance is considered with p values < 0.05. In the case of detecting overall differences, post-hoc comparisons using Dunn’s test with Bonferroni adjustments were planned between each group. The effect of group assignment on the change in the knowledge outcomes repeated at 2 weeks was assessed using linear regression, adjusting for baseline measurement. Intervention group was not masked prior to this planned analysis.

Effect size and precision (95% confidence interval) for non-parametric tests were estimated using η² (rule of thumb for small effect 0.01; medium = 0.06; large = 0.14).^38^ Sensitivity analyses investigated the robustness of primary analyses for participants who passed the attention check. Stratified analyses explored differences according to key characteristics: gender (man; woman), age (≤60 years vs >60 years), education (university education vs no university education), chronic disease (no chronic disease; at least one self-reported chronic disease), and previous ChatGPT use (none in the past 6 months; at least a few times in the past 6 months). Stratified analyses for health literacy and digital health literacy were not explored given the relatively small number in some cells. A further sensitivity analysis was performed by imputing the primary outcomes using multiple imputation using chained equations for those who did not complete the survey.

As a low-risk study, we did not anticipate any serious harms.

### Sample Size

The total sample size required was 2000 participants. With a 2 x 5 parallel group design, with 90%, alpha = 0.05, and a small effect size, we estimated 160 participants per intervention group. A further 20% was added to account for drop-out at follow-up.

### Patient and public involvement

A working group comprising nine people helped shape the intervention designs. After describing the goals of the project and providing foundational information about generative AI, working group members discussed their needs, priorities, and expectations for the tool. Insights from these discussions were incorporated into four prototypes to elicit specific feedback in a second workshop. Design strategies that the working group and healthdirect staff considered essential were incorporated into the AI-enhanced format. For more detail see Appendix C.

## RESULTS

### Sample characteristics

2546 participants were randomised to an intervention group, of which 2062 comprised the analysis sample (Figure 1; Tables 1 and 2). Across the sample 49% of participants identified as man or male, 51% identified as woman or female, and the median age was 49 years (IQR=28). Fifty-six per cent of participants had obtained a university degree and 16% were identified as having limited health literacy based on the screening item. Two-fifths (43%) of the sample had heard of healthdirect, and 14% had previously used the healthdirect symptom checker tool. The sample was familiar with generative AI tools like ChatGPT, with 56% of participants reporting that they had used ChatGPT at least a few times in the past 6 months (n=1164), and 36% reported that they had used it for health advice (n=740). The most common reason for using ChatGPT for health purposes was to ask about symptoms (19%, n=385). The median digital health literacy score was 31.0 (IQR: 5.0), indicating moderate to high confidence in this domain. No harms were reported in this study. Characteristics of participants who completed follow-up (N=1531, 74.2%), and were similar to those of the baseline sample (eTable 1).

**Table 1.** Participant characteristics, low acuity symptom level (self-care), by format intervention group (n, %, unless otherwise specified)

| Variable | Standard<br>(original tool)<br>N = 215<sup>1</sup> | AI-enhanced<br>N = 206<sup>1</sup> | AI-enhanced +<br>Multimedia<br>N = 202<sup>1</sup> | AI-enhanced +<br>Numbered<br>steps<br>N = 206<sup>1</sup> | AI-enhanced +<br>Detailed info<br>about AI<br>N = 212<sup>1</sup> |
| --- | --- | --- | --- | --- | --- |
| **Age (years)** <sup>1</sup> | 49.0 (35.0, 64.0) | 47.5 (34.0, 63.0) | 49.0 (35.0, 63.0) | 51.0 (39.0, 65.0) | 48.5 (36.0, 63.0) |
| **Gender** |  |  |  |  |  |
| Man | 98 (46%) | 97 (47%) | 96 (48%) | 100 (49%) | 110 (52%) |
| Woman | 117 (54%) | 109 (53%) | 105 (52%) | 105 (51%) | 101 (48%) |
| Trans and/or gender diverse | 0 (0%) | 0 (0%) | 0 (0%) | 1 (0.5%) | 1 (0.5%) |
| I use a different term / Prefer<br>not to say | 0 (0%) | 0 (0%) | 1 (0.5%) | 0 (0%) | 0 (0%) |
| **Born in Australia** | 157 (73%) | 162 (79%) | 161 (80%) | 164 (80%) | 152 (72%) |
| **What is the main language you**<br>**speak at home?** |  |  |  |  |  |
| Other | 19 (8.8%) | 16 (7.8%) | 15 (7.4%) | 13 (6.3%) | 17 (8.0%) |
| English | 196 (91%) | 190 (92%) | 187 (93%) | 193 (94%) | 195 (92%) |
| **Are you of Aboriginal or Torres**<br>**Strait Islander origin?** |  |  |  |  |  |
| Aboriginal and/or Torres Strait<br>Islander | 6 (2.8%) | 9 (4.4%) | 6 (3.0%) | 6 (2.9%) | 7 (3.3%) |
| No | 208 (97%) | 196 (96%) | 196 (97%) | 199 (97%) | 202 (97%) |
| Unknown | 1 (0.5%) | 1 (0.5%) | 0 (0%) | 1 (0.5%) | 3 (1.4%) |
| **Highest level of education** |  |  |  |  |  |
| No school or other<br>qualifications | 0 (0%) | 3 (1.5%) | 2 (1.0%) | 5 (2.4%) | 2 (0.9%) |
| School Certificate or<br>intermediate certificate (or<br>equivalent) | 11 (5.1%) | 10 (4.9%) | 6 (3.0%) | 8 (3.9%) | 17 (8.0%) |
| Higher school certificate or<br>leaving certificate (or equivalent) | 30 (14%) | 38 (18%) | 28 (14%) | 39 (19%) | 30 (14%) |
| Trade Apprenticeship | 10 (4.7%) | 7 (3.4%) | 10 (5.0%) | 7 (3.4%) | 4 (1.9%) |
| Diploma or Certificate | 41 (19%) | 27 (13%) | 40 (20%) | 34 (17%) | 33 (16%) |
| University degree | 123 (57%) | 121 (59%) | 116 (57%) | 113 (55%) | 126 (59%) |

| Variable | Standard<br>(original tool)<br>N = 215<sup>†</sup> | AI-enhanced<br>N = 206<sup>†</sup> | AI-enhanced +<br>Multimedia<br>N = 202<sup>†</sup> | AI-enhanced +<br>Numbered<br>steps<br>N = 206<sup>†</sup> | AI-enhanced +<br>Detailed info<br>about AI<br>N = 212<sup>†</sup> |
| --- | --- | --- | --- | --- | --- |
| **Health literacy screener** |  |  |  |  |  |
| Adequate health literacy | 181 (84%) | 166 (81%) | 178 (88%) | 178 (86%) | 176 (83%) |
| Limited health literacy | 34 (16%) | 40 (19%) | 24 (12%) | 28 (14%) | 36 (17%) |
| **eHEALS Score (8-40)**<sup>†</sup> | 31.0 (28.0, 33.0) | 31.0 (28.0, 33.0) | 31.0 (28.0, 33.0) | 32.0 (28.0, 33.0) | 31.0 (27.0, 34.0) |
| **Number of chronic conditions** |  |  |  |  |  |
| 0 | 119 (55%) | 117 (57%) | 118 (58%) | 103 (50%) | 114 (54%) |
| 1 | 63 (29%) | 56 (27%) | 68 (34%) | 62 (30%) | 68 (32%) |
| 2 | 27 (13%) | 24 (12%) | 11 (5.4%) | 30 (15%) | 19 (9.0%) |
| 3+ | 6 (2.8%) | 9 (4.4%) | 5 (2.5%) | 11 (5.3%) | 11 (5.2%) |
| **healthdirect use** |  |  |  |  |  |
| Helpline | 25 (12%) | 24 (12%) | 22 (11%) | 20 (9.7%) | 24 (11%) |
| Symptom Checker | 32 (15%) | 31 (15%) | 26 (13%) | 35 (17%) | 27 (13%) |
| Web-based information | 43 (20%) | 46 (22%) | 51 (25%) | 42 (20%) | 40 (19%) |
| Videos | 7 (3.3%) | 7 (3.4%) | 13 (6.4%) | 8 (3.9%) | 10 (4.7%) |
| Service Finder | 16 (7.4) | 23 (11%) | 14 (6.9%) | 18 (8.7%) | 19 (9.0%) |
| Mobile App | 11 (5.1%) | 17 (8.3%) | 10 (5.0%) | 12 (5.8%) | 16 (7.5%) |
| **ChatGPT use for health information (in the last 6 months)** |  |  |  |  |  |
| A few times | 34 (16%) | 56 (27%) | 35 (17%) | 37 (18%) | 49 (23%) |
| Once a month | 14 (6.5%) | 11 (5.3%) | 16 (7.9%) | 12 (5.8%) | 11 (5.2%) |
| Once a week | 9 (4.2%) | 9 (4.4%) | 8 (4.0%) | 9 (4.4%) | 14 (6.6%) |
| More than once a week | 8 (3.7%) | 12 (5.8%) | 7 (3.5%) | 8 (3.9%) | 10 (4.7%) |
| Has not used ChatGPT for health | 150 (70%) | 118 (57%) | 136 (67%) | 140 (68%) | 128 (60%) |

**Table 2.**
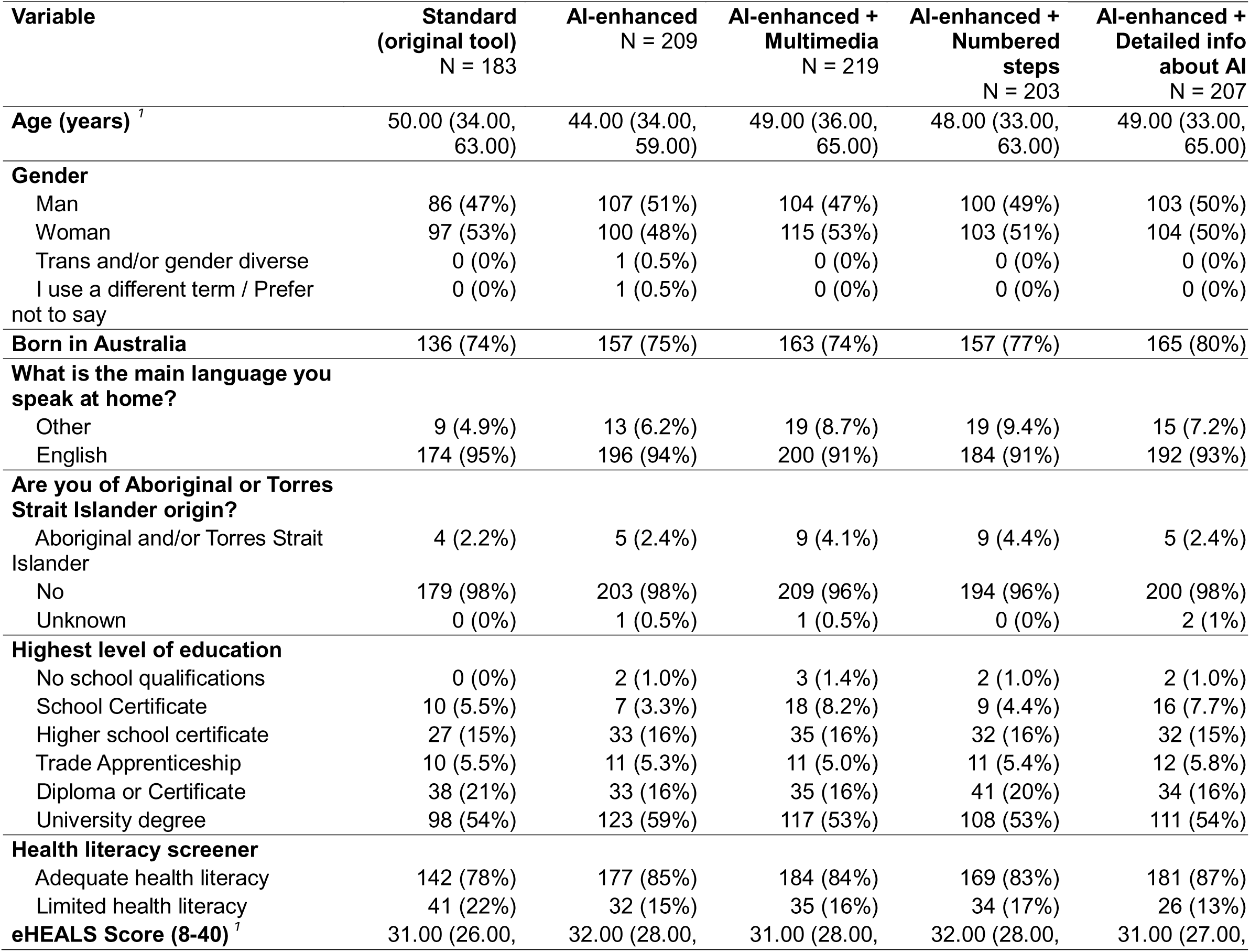
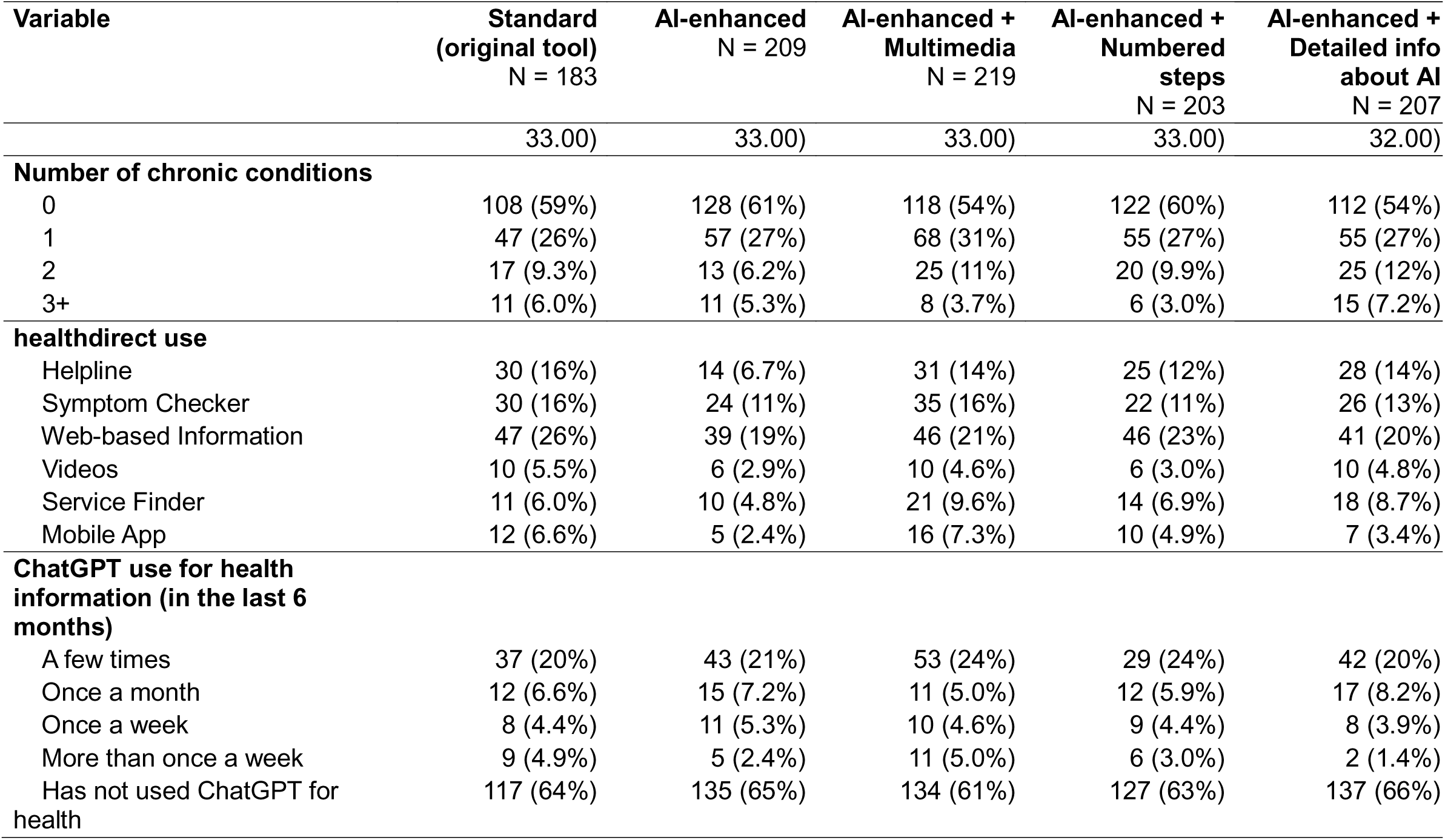
Participant characteristics, moderate acuity symptom level (see a GP in 24 hours), by format intervention group (n, %, unless otherwise specified)

### Evaluation of Symptom Checker intervention formats

Descriptive statistics for outcomes are shown in Figure 3 and eTables 2 and 3.

**Figure 3.**
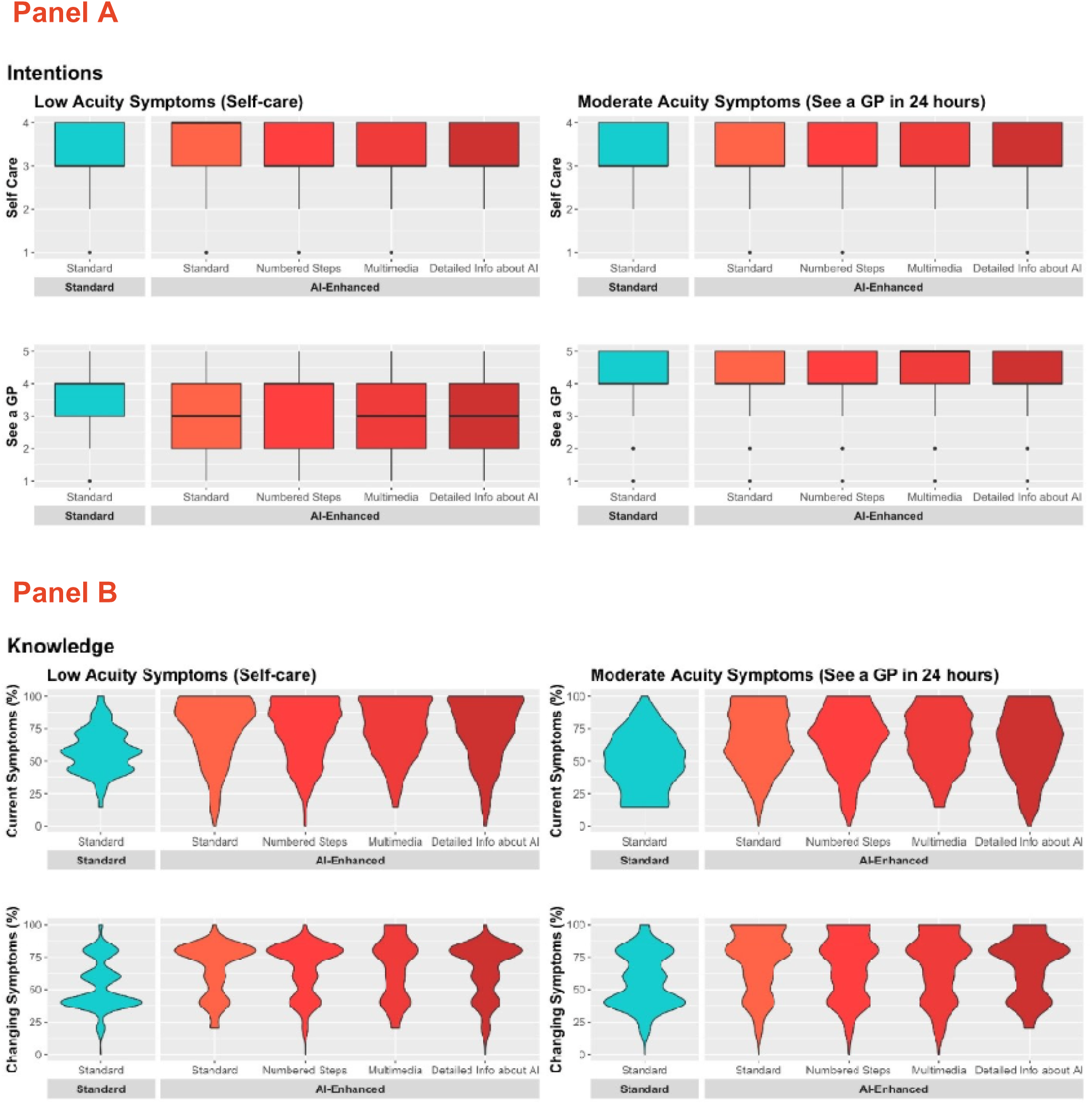
Distribution of intention and knowledge scores (immediately post-intervention), by intervention group and symptom acuity level.

#### Low acuity symptoms (self-care)

##### Intention

For the low acuity presentation, which advised to self-care at home, there was a statistically significant effect of intervention format on intention to see a GP in 24 hours (H(4) = 13.42, p = 0.009). The effect size was small (η²= 0.009; 95% CI:0.001-0.03). Post-hoc analysis showed that participants who viewed the AI enhanced format (median = 3.00) had significantly lower intentions to see a GP within 24 hours than those in the Standard (original) tool group (median = 4.00; adjusted p = 0.003). There was no significant effect of intervention format on intention to follow self-care advice for the low acuity symptoms (H(4) = 1.41, p=0.843).

##### Knowledge

For the low acuity symptoms there was a statistically significant effect of intervention format on both knowledge measures – managing current symptoms and managing changing symptoms (H(4) = 108.16, p <0.001; H(4) = 67.38, p <0.001, respectively). The effect size was moderate for both outcomes (current symptoms: η²= 0.101; 95% CI:0.07-0.14; managing symptoms: η²= 0.061; 95% CI:0.04-0.10). Post-hoc analyses demonstrated that mean ranks for all AI-enhanced groups were significantly higher than those of the Standard group (original tool) (all adjusted p’s <0.001; Figure 3 and eTable 2).

##### Trust

Intervention format did not have a significant effect on any trust outcomes for the low acuity symptoms (eTable 2).

##### Acceptability

Intervention format did not have a significant effect on acceptability for the low acuity symptoms (eTable 2).

#### Moderate acuity symptoms (see a GP in 24 hours)

##### Intention

For the moderate acuity symptoms, there was no significant effect of intervention format on intention to see a GP in 24 hours (H(4) = 8.35, p=0.080) or to follow the self-care advice (H(4) = 2.03, p=0.730) (eTable 3).

##### Knowledge

For the moderate acuity symptoms there was a statistically significant effect of intervention format on both knowledge measures – managing current symptoms and managing changing symptoms (H(4) = 72.14, p <0.001; H(4) 52.93, p <0.001, respectively). The effect size was moderate for both outcomes (current symptoms: η² = 0.067; 95% CI:0.04-0.14; managing symptoms: η²= 0.048; 95% CI(0.03-0.08)). Post-hoc analyses demonstrated that mean ranks for all AI-enhanced groups were significantly higher than those of the Standard (original tool) group (all adjusted p’s <0.001; eTable 3).

##### Trust

Intervention format did not have a significant effect on any trust outcomes for the moderate acuity symptoms (eTable 3).

##### Acceptability

Intervention format did not have a significant effect on acceptability for the moderate acuity symptoms (eTable 3).

#### Knowledge at follow-up

Within each symptom acuity level, all formats reported similar levels of knowledge about current and changing symptoms at two-week follow-up (time point 2) (eTables 4-5).

For the low acuity symptoms, the changes between time point 1 and time point 2 represented a small but significantly larger decrease in knowledge of current symptoms for participants in the AI-enhanced group, compared to those in the Standard group (original tool) (MD= −14.3%, 95% CI= −18.3% to −12.3%, p=0.006). For the moderate acuity symptoms (see a GP in 24 hours), we observed that two of the AI-enhanced formats showed small but significantly larger decreases in knowledge of current symptoms relative to the Standard group (original tool) (Detailed info about AI: MD= −8.0%, 95% CI= −11.1% to −4.9%, p=0.023); Multimedia: MD= −9.4%, 95% CI= −12.3% to −6.6%, p=0.003). There were no significant changes in knowledge about changing symptoms over time.

#### Exploratory analyses

Participants who correctly recalled at least one of the two symptoms in the health scenario (vomiting, fever), and no incorrect symptoms, were included in a sensitivity analysis to explore the impact of attention on the statistical findings. Some participants (9.3% of the total sample, n=191) did not demonstrate appropriate attention. Analyses following the removal of inattentive participants, yielded very similar results, demonstrating that study findings were robust to inattentive participants (eTables 6 and 7).

Stratified analyses reported some differences in the effects of the Standard (original tool) and pooled effects of the AI-enhanced formats, depending on participant characteristics. These differences were only observed for the low acuity symptoms, in which the symptom checker recommended that people self-care at home: participants who viewed any AI format reported lower intentions to see a GP in 24 hours than those in the Standard (original tool) group if they had a university education (p=0.012), had not used ChatGPT in the past 6 months (p=0.012), did not have a chronic condition (p=0.007), and were ≤60 years old (p=0.008). This pattern was not observed for people without a university education (p=0.12), those who had used ChatGPT at least a few times in the past 6 months (p=0.10), people who reported at least one chronic condition (p=0.2), or those who were more than 60 years old (p=0.2). No differences were observed for gender.

Characteristics of those who did not complete the survey were similar overall and within intervention groups, to those included in the analysis. The sensitivity analysis using multiple imputation of the missing primary outcomes showed comparable results (eTable 8).

## DISCUSSION

This study evaluated the effects of personalised, AI-enhanced online symptom checker formats on intentions to seek care, knowledge about managing symptoms, and trust and acceptability of the tool. For low acuity symptoms, where advice was to self-care, tailored formats reduced intentions to seek potentially unnecessary professional care. For moderate acuity symptoms, where advice was to see a GP in 24 hours, there was no effect of format on intentions to follow this advice. Across both symptom acuity levels, there was no impact of format on intentions to self-care. More tailored formats improved knowledge for both symptom acuity levels compared to existing formats at immediate follow up, however, these gains were not sustained at two weeks. Formats that included explicit statements about AI did not significantly affect participants’ high levels of trust in the advice or acceptability of the tool.

Although several studies have investigated users’ intentions to follow symptom checker advice,^11^ ^13–15^ few have quantitatively compared the effects of different formats. Our findings suggest that for low acuity symptoms, more personalised information may reduce potentially unnecessary use of health services. Two key design features may have contributed to this effect: the use of actionable plain language advice for managing both current and changing symptoms; and a clearer description of the rationale for managing the symptoms at home. These features address well-documented issues for symptom checkers.^5^ ^10^ ^17^ ^18^ ^20^ ^21^

This is also one of the first studies to explore public responses to the use of generative AI in online symptom checkers. Explicit statements about the use of AI did not significantly impact on trust or acceptability of the tool. This finding is consistent with the broader literature. People are increasingly turning to general purpose AI tools such as ChatGPT for health advice,^7–9^ and as people become more competent and familiar with generative AI tools, they are likely to also become more trusting^39^ (although there is also evidence that those most familiar are also some of the least trusting).^40^ In this study the AI was also integrated into a recognisable national health service (healthdirect), with visual cues such as government logos and URLs, and a statement about clinician review of the content. Both features may have enhanced perceived reliability.^41^ ^42^

The strengths of this study are the use of pre-registered randomised controlled-trial study design to enhance internal validity of the findings, and community involvement to improve acceptability of the tool to people in the community. The study also has several limitations. We observed high intentions across all groups, potentially limiting our capacity to detect differential effects of the formats. The primary outcome (intention) and knowledge were purpose-built items. Though highly relevant to the study, these assessments were not validated, and the items may not reliably and accurately reflect the underlying constructs. This is a common challenge, particularly for performance-based assessment of knowledge. Lastly, the symptom checker formats were presented as screenshots. Participants may have responded differently to interactive versions, particularly those with a stronger emphasis on visual design e.g. the multimedia format.

Further research is needed to improve the external validity and generalisability of the findings. For example, enabling participants to engage with interactive versions of the tool, rather than screenshots, would deliver stronger evidence about the impact of different visual design elements on the outcomes of interest. This study used a hypothetical scenario; allowing participants to respond to real concerns would provide more generalisable findings across diverse health contexts and levels of acuity.

## Conclusion

Symptom checkers are widely available worldwide to support people to make informed decisions about when to seek advice from healthcare services and how to manage symptoms at home. This study found that people were better equipped with knowledge to manage symptoms when they viewed online symptom checker formats that gave more tailored advice. There was also some evidence that these more tailored formats may help reduce the potentially unnecessary use of primary care services for symptoms that can be managed safely at home. The explicit use of AI did not impact significantly on trust or acceptability of online symptom checker. Future research should investigate these formats using interactive prototypes across a wider variety of health contexts.

## Supporting information

CONSORT checklist

## ACKNOWLEDGEMENTS

We would like to acknowledge Ms Momo Hudson-Barton for her support in preparing the figures used in this manuscript. We would also like to acknowledge the contributions of our workshop members Geoffrey Edlund, Ivan CK Ma, Trang Vu, Sharon Ng, Waren Nadesan, Gauri Kapoor, Cheryl Knight, and Lorna Butters. We would also like to acknowledge Rochelle Seneviratna support in managing the project.

## FUNDING

Julie Ayre and Kirsten McCaffery are supported by National Health and Medical Research Council fellowships (APP2017278, APP2016719). This work was funded by a research contract awarded to the Sydney Health Literacy Lab, University of Sydney with healthdirect, a government funded entity that provides national evidence-based health information and advice in Australia (#CON230550-Symptom Checker Gen AI Project-healthdirect).

## COMPETING INTERESTS

Karen Gallagher, Angelica Scott, Alison Woods, Christopher Ng, and Yuanee Wickramasinghe are employed by healthdirect.

## DATA AVAILABILITY

De-identified data produced in the present study can be made available upon request.

## Notes

### Clinical Trial

Australian New Zealand Clinical Trials Registry (ACTRN12625000474459p)

### Author Declarations

approved by the University of Sydney Research Ethics Committee (2025/HE000143).

### Summary of Updates

The consort flow diagram (Figure 1) was corrected with additional reasons as to why participants were excluded from analysis. There is now additional text and appendix material (eTable 8 and associated in-text results) relating to a sensitivity analysis that included participants who did not complete the survey.

