## Appendix for "Evaluation of symptom checker formats to support health literacy and trust in AI: Results from an online randomised-controlled trial"

### Appendix A: Intervention groups

Note: For viewing ease we present eFigures 2-11 on one page. Participants in the trial scrolled to view each format in its entirety.

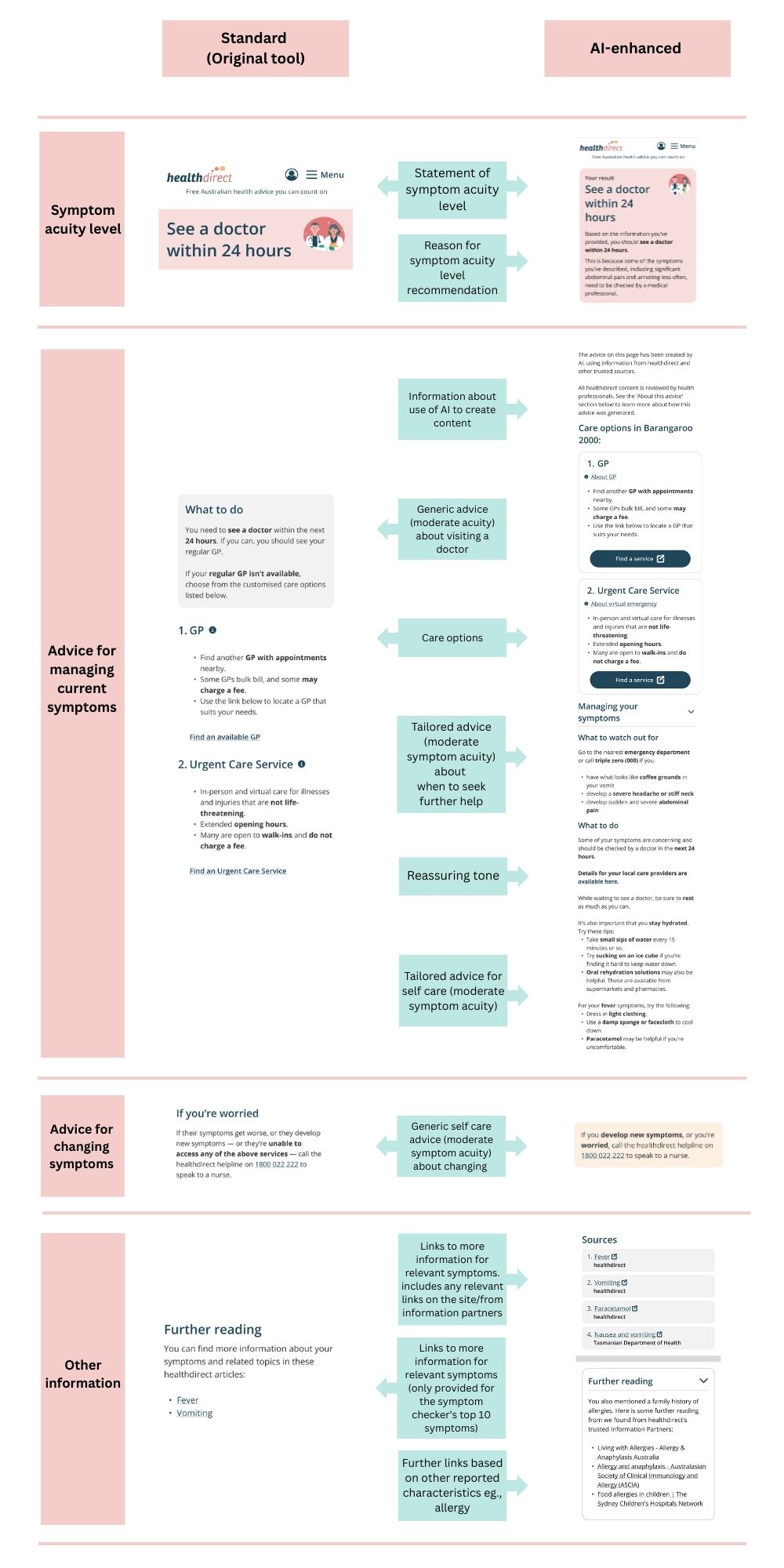

**eFigure 1. Comparison of standard and AI-enhanced formats, moderate acuity symptoms**

**eFigure 2. Low acuity symptoms, Standard (original tool)**

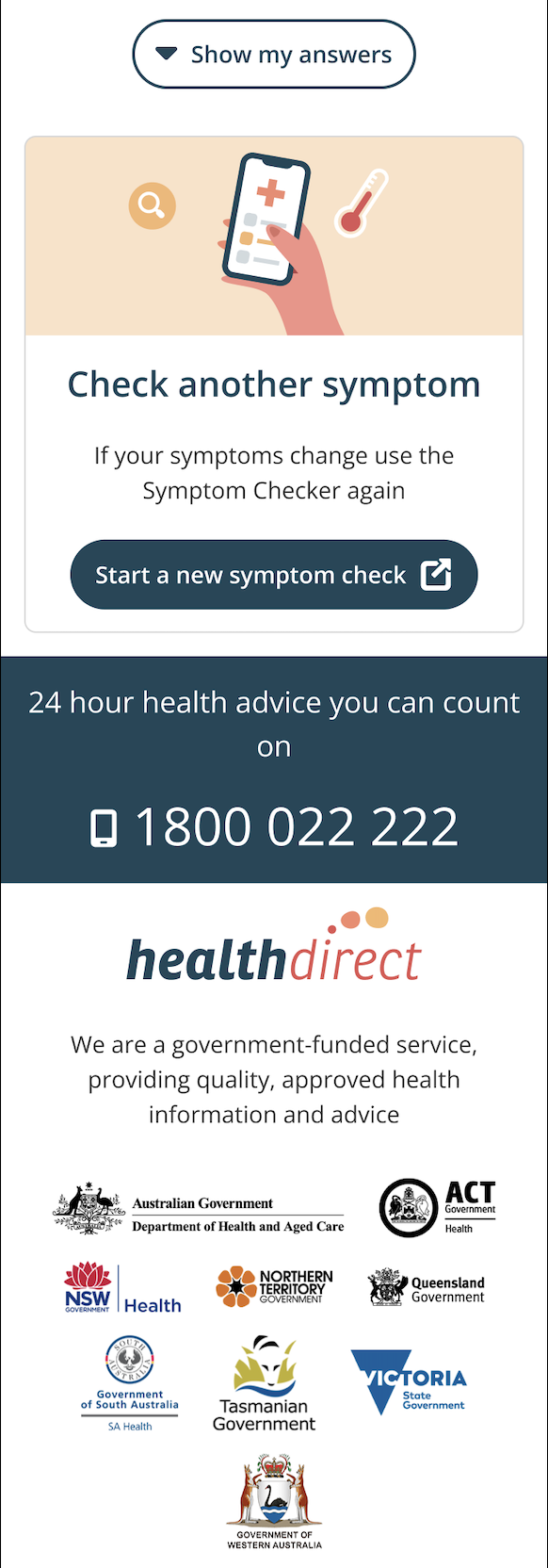

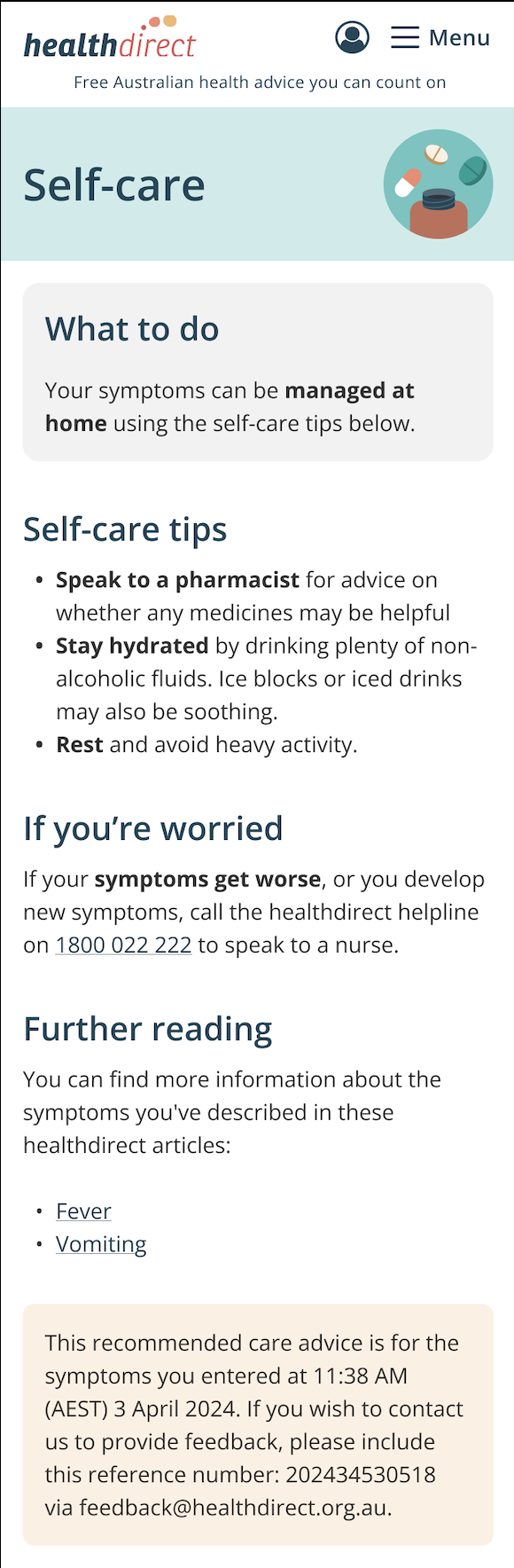

**eFigure 3. Moderate acuity symptoms, Standard (original tool)**

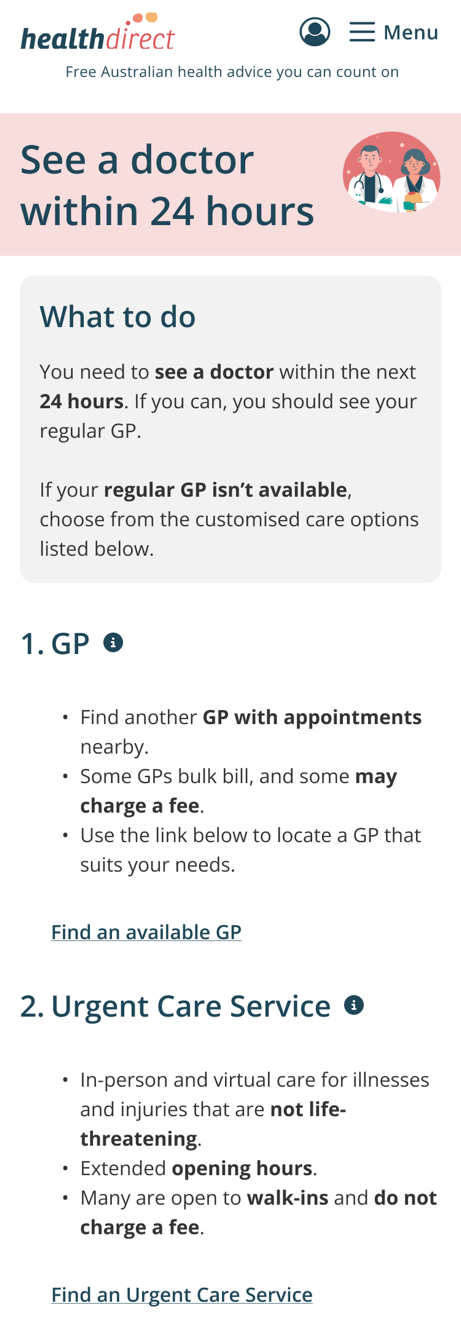

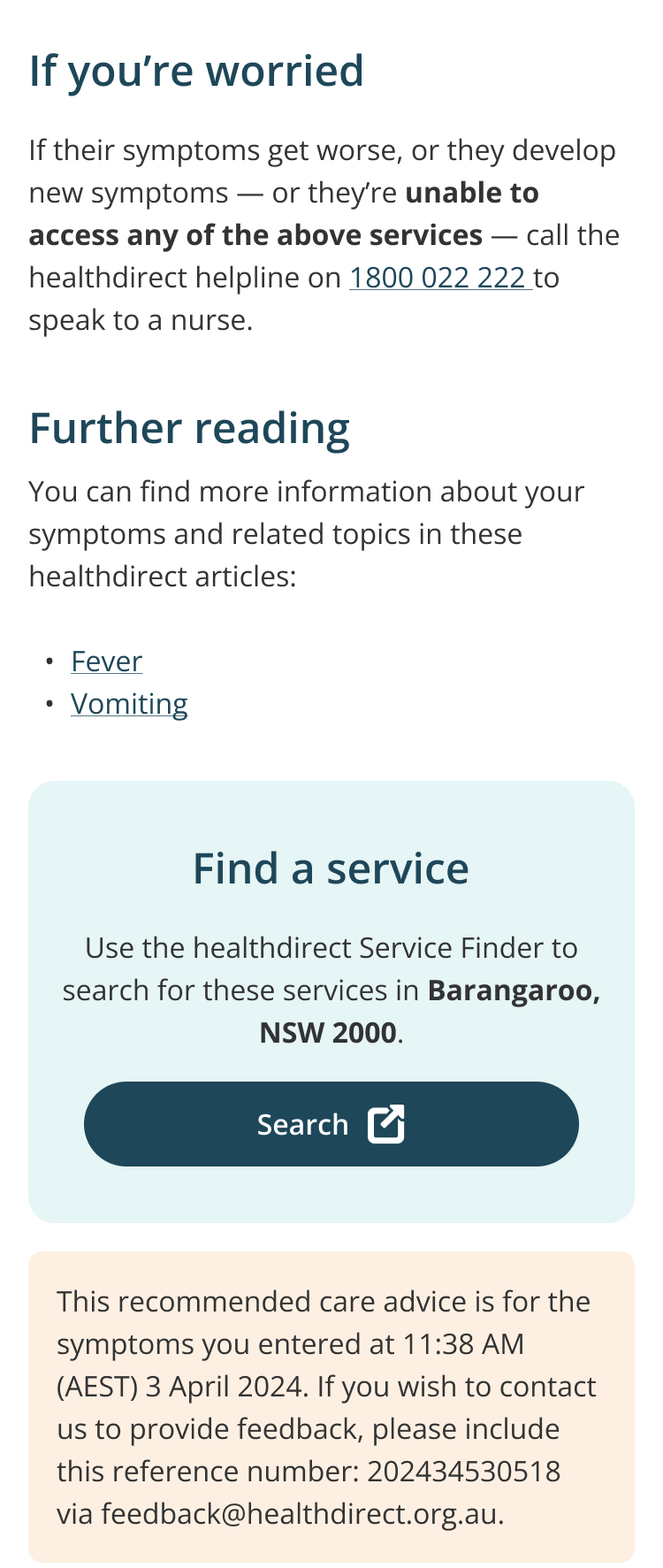

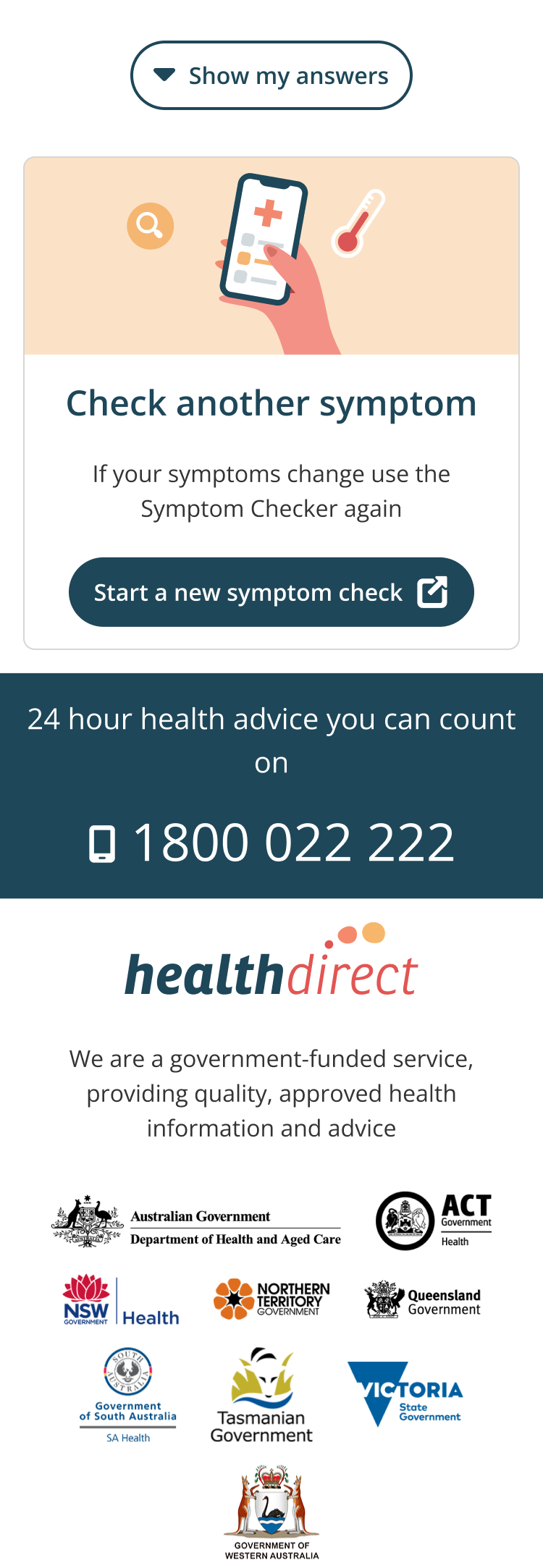

**eFigure 4. Low acuity symptoms, AI-enhanced**

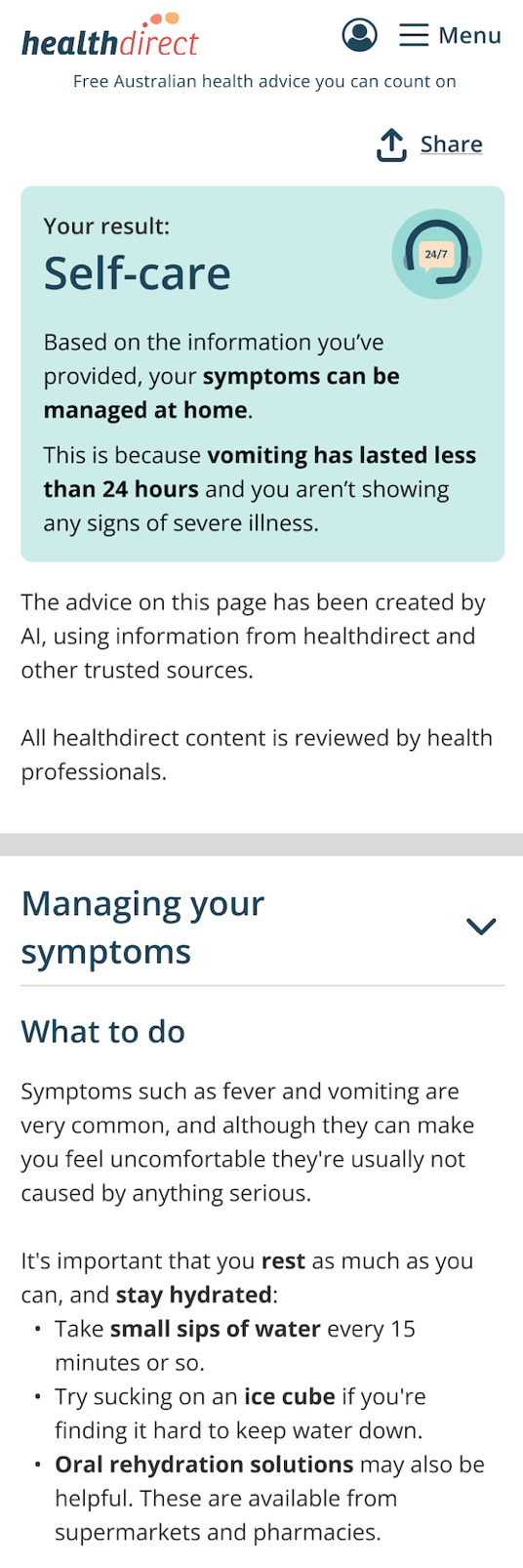

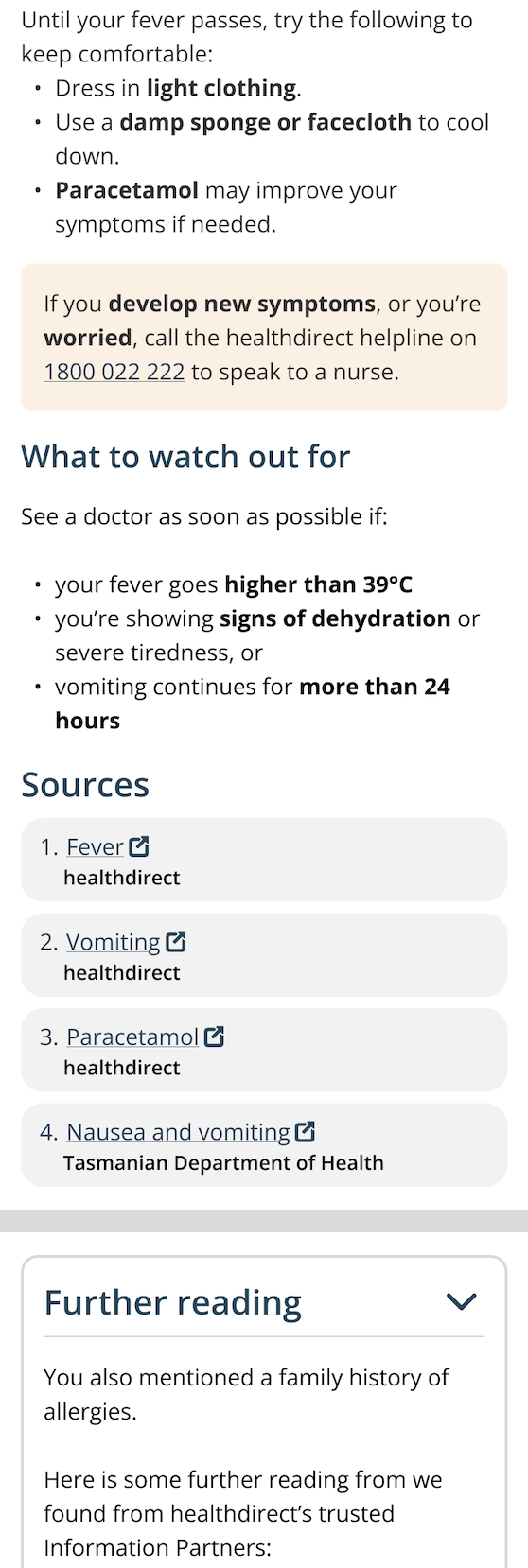

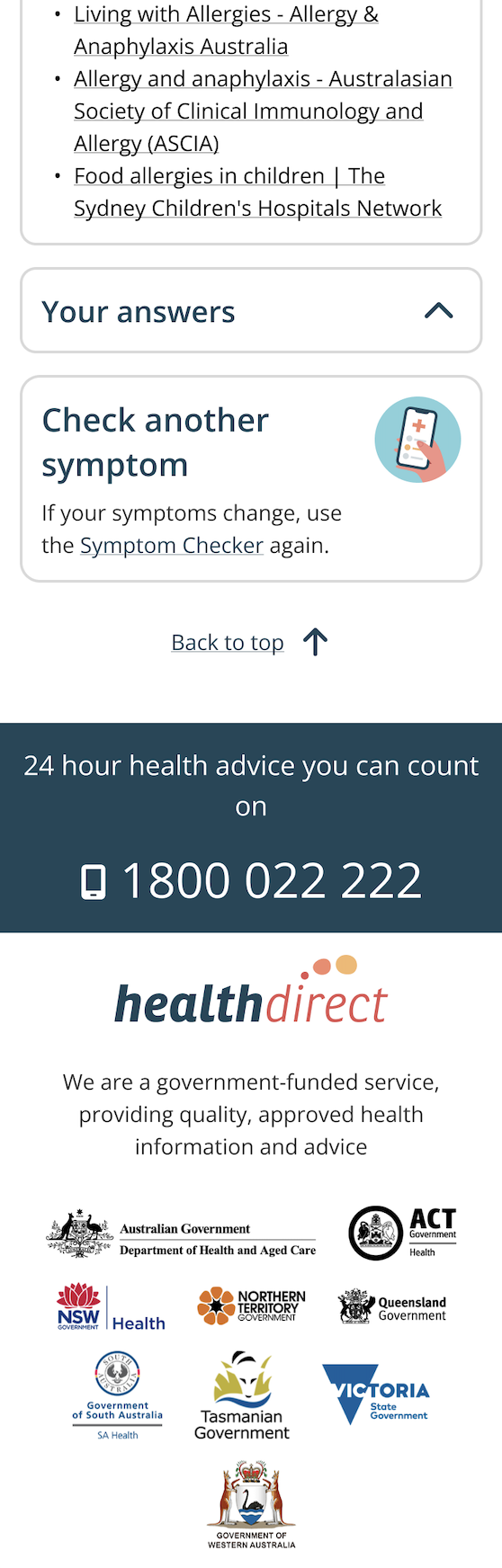

**eFigure 5. Moderate acuity symptoms, AI-enhanced**

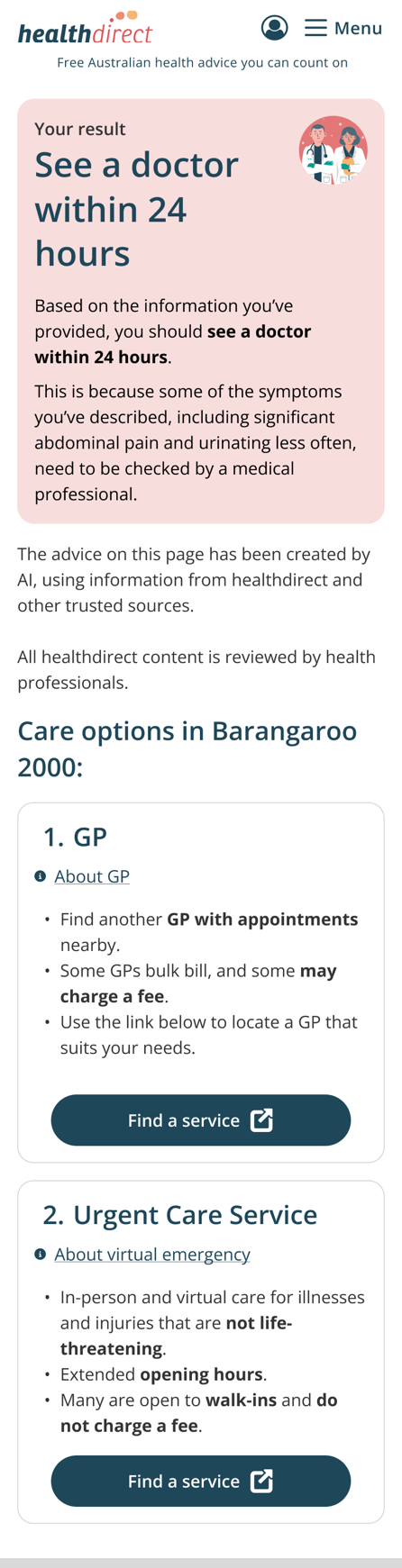

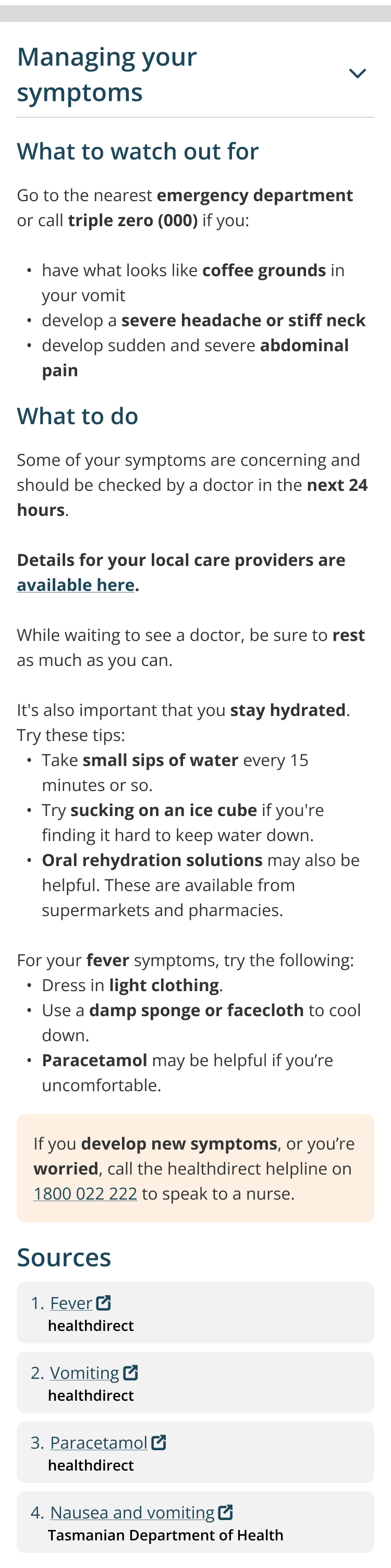

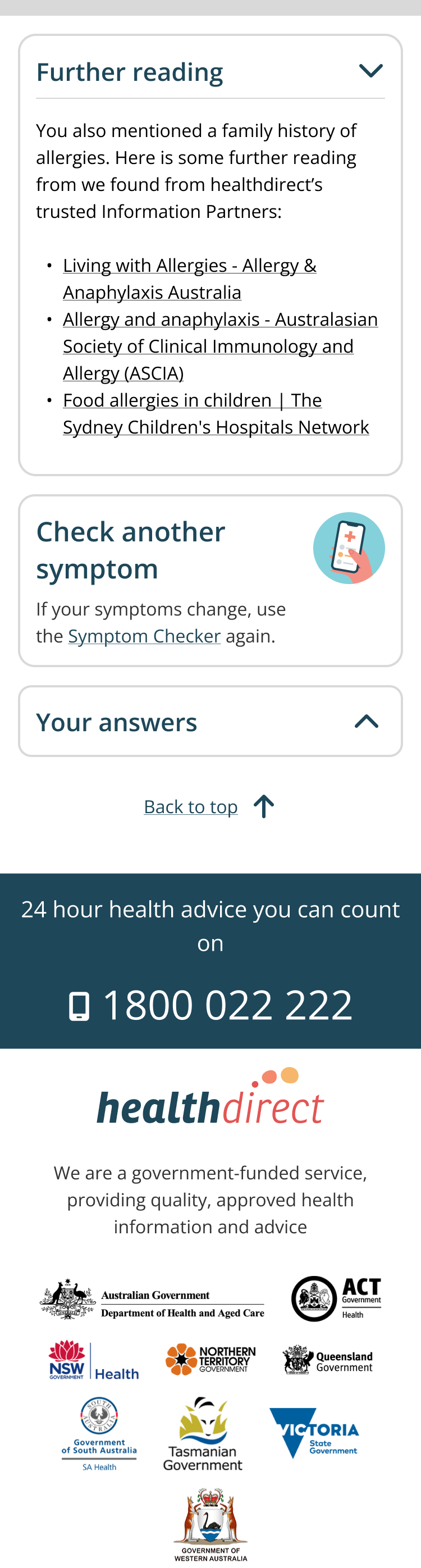

**eFigure 6. Low acuity advice, AI-enhanced + numbered steps**

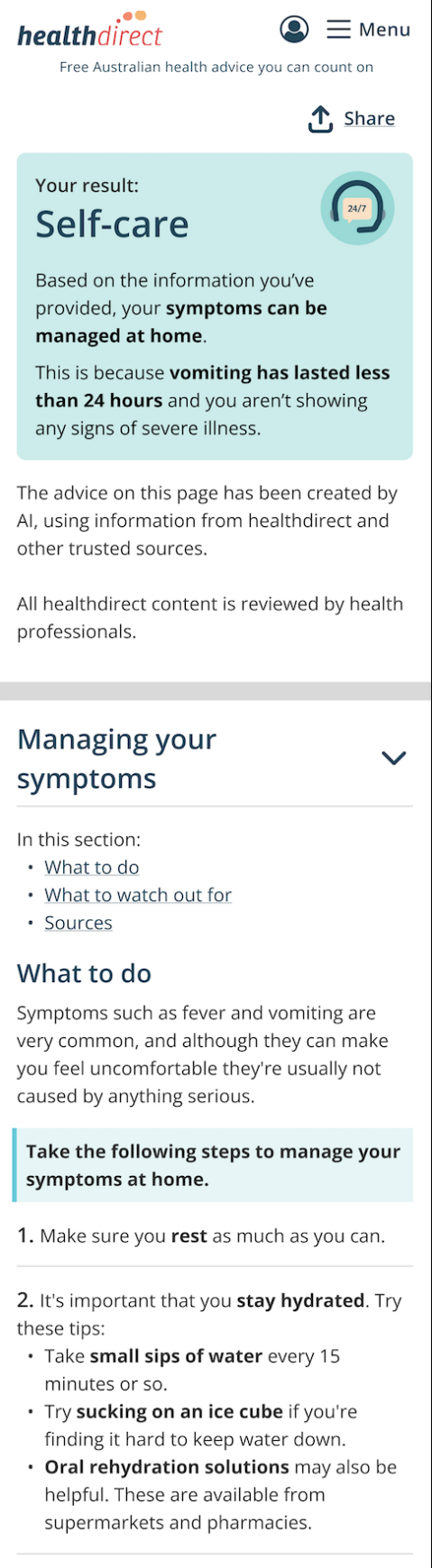

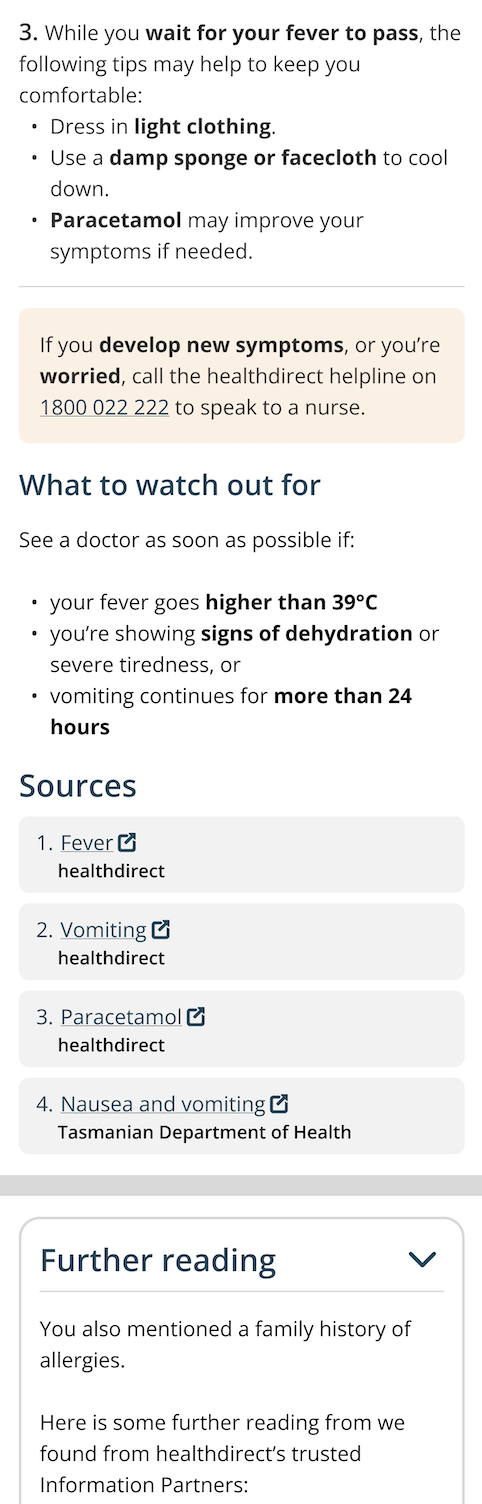

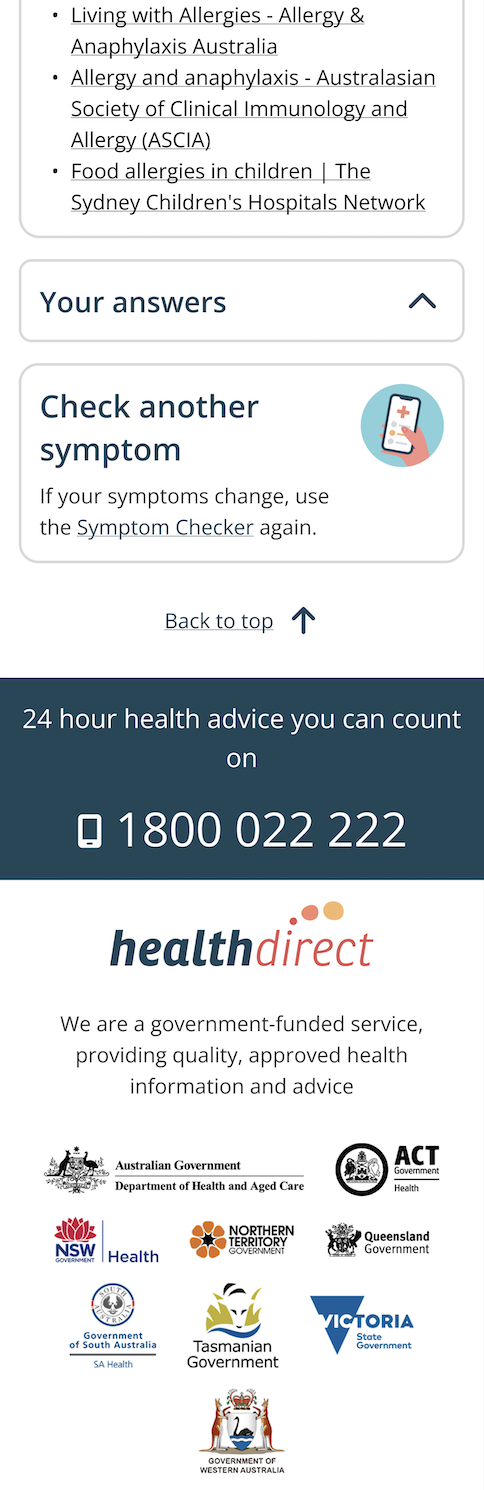

**eFigure 7. Moderate acuity symptoms, AI-enhanced + numbered steps**

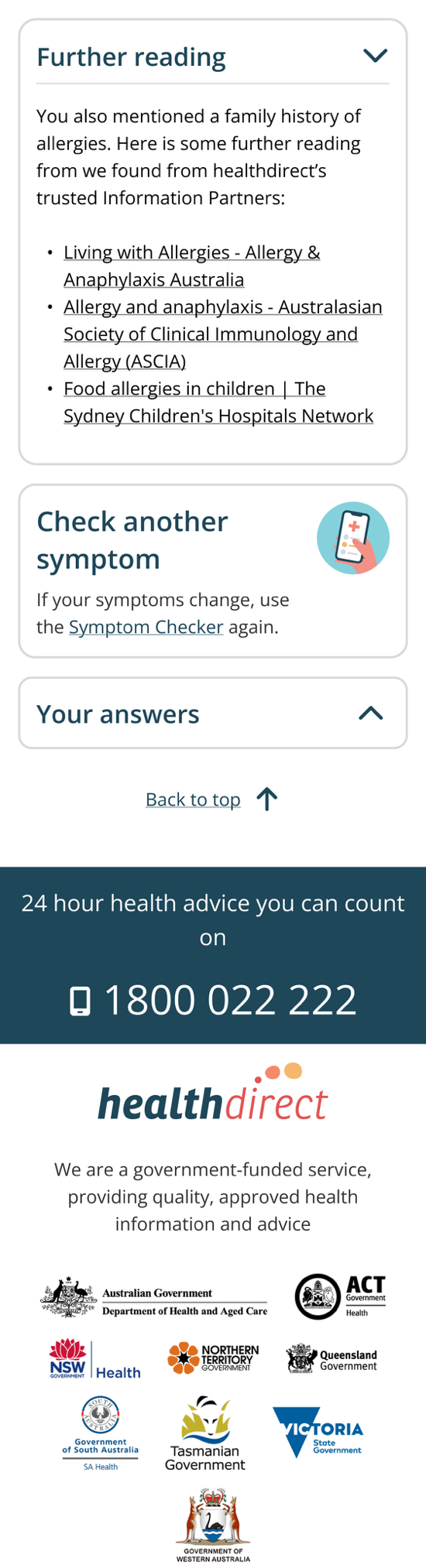

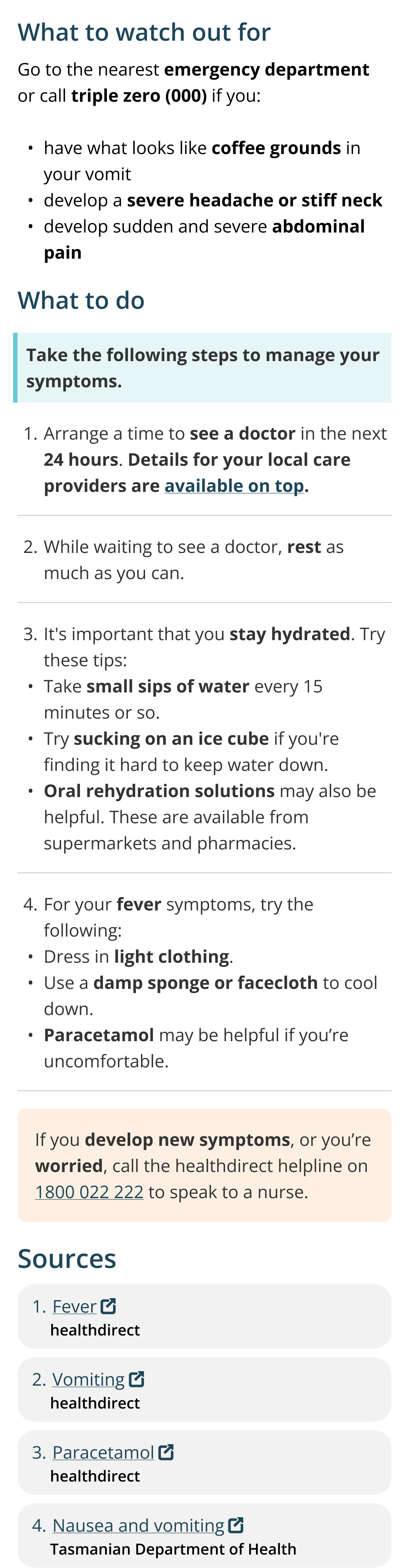

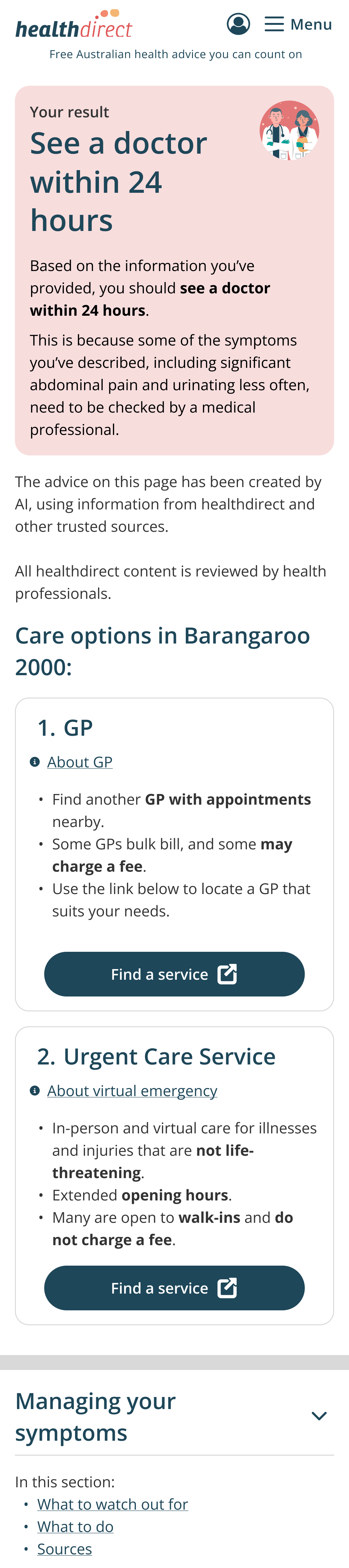

**eFigure 8. Low acuity symptoms, AI-enhanced + multimedia**

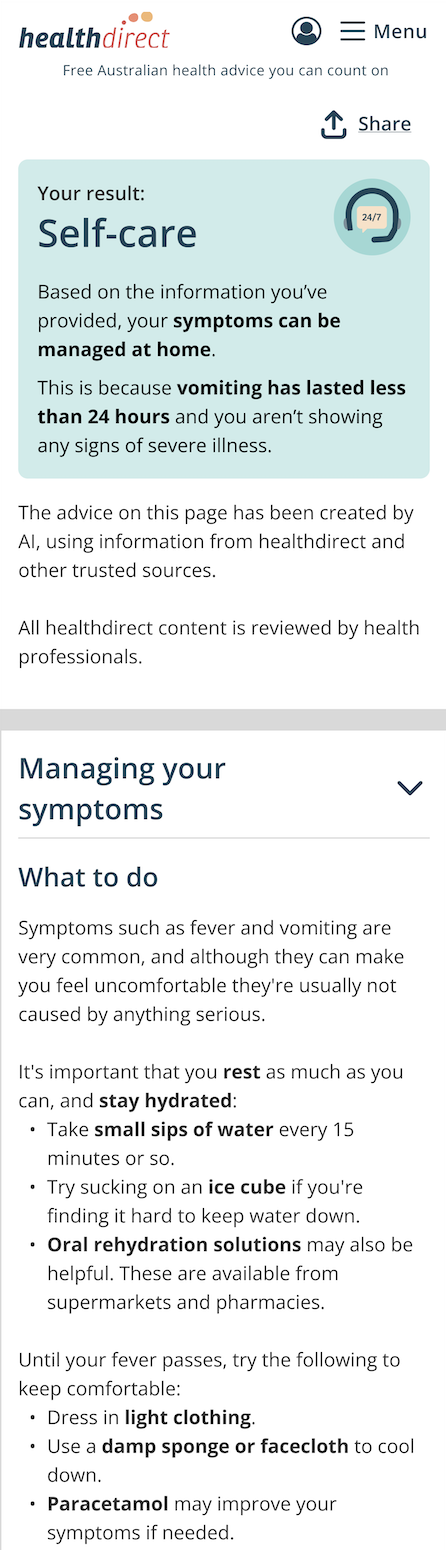

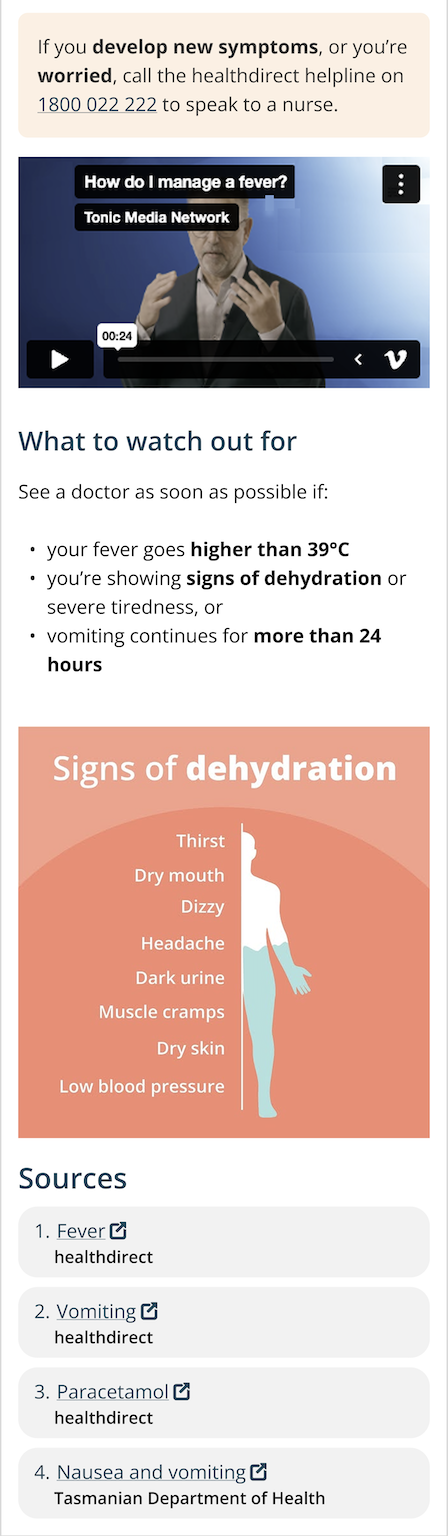

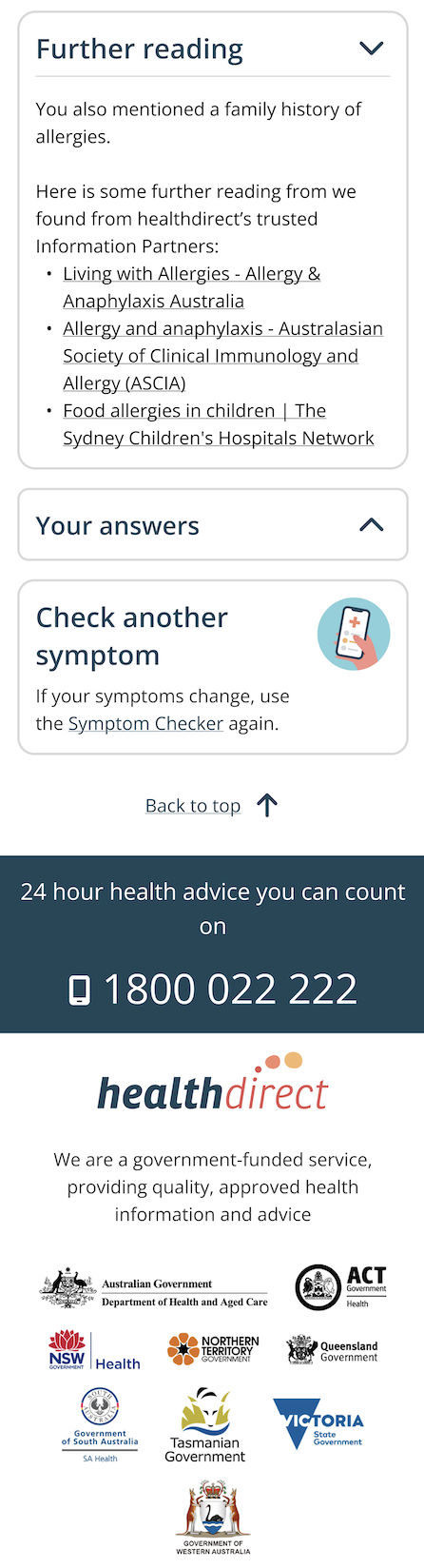

**eFigure 9. Moderate acuity symptoms, AI-enhanced + multimedia**

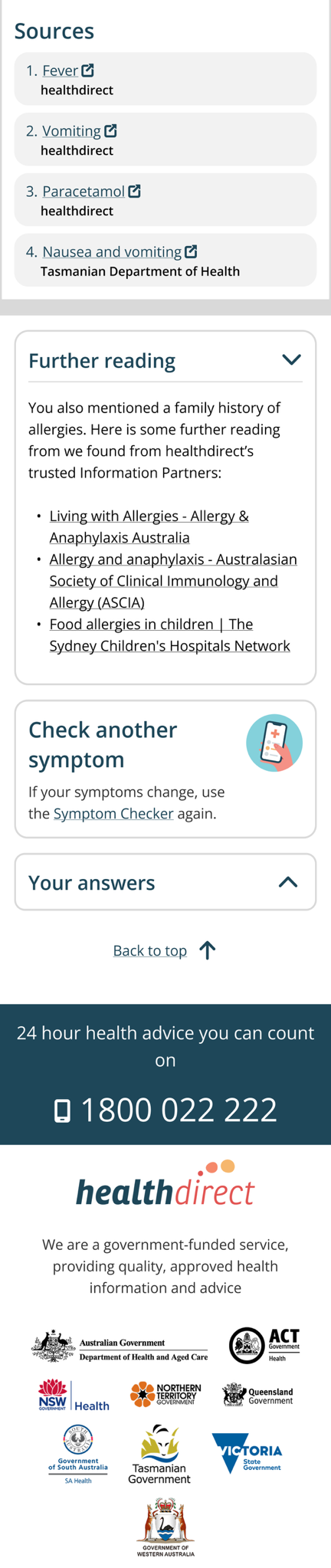

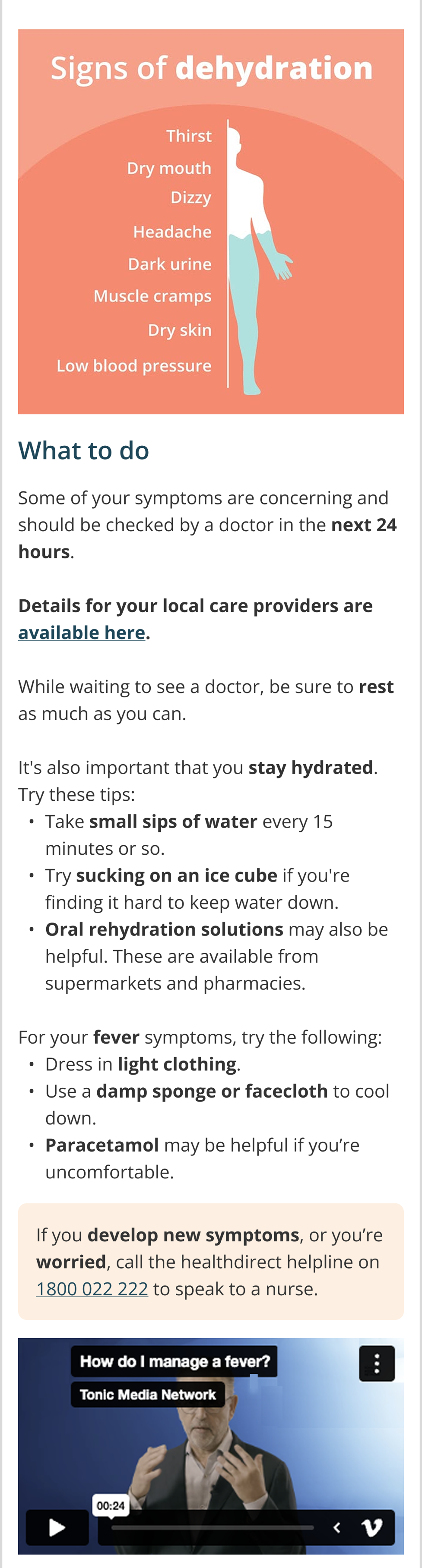

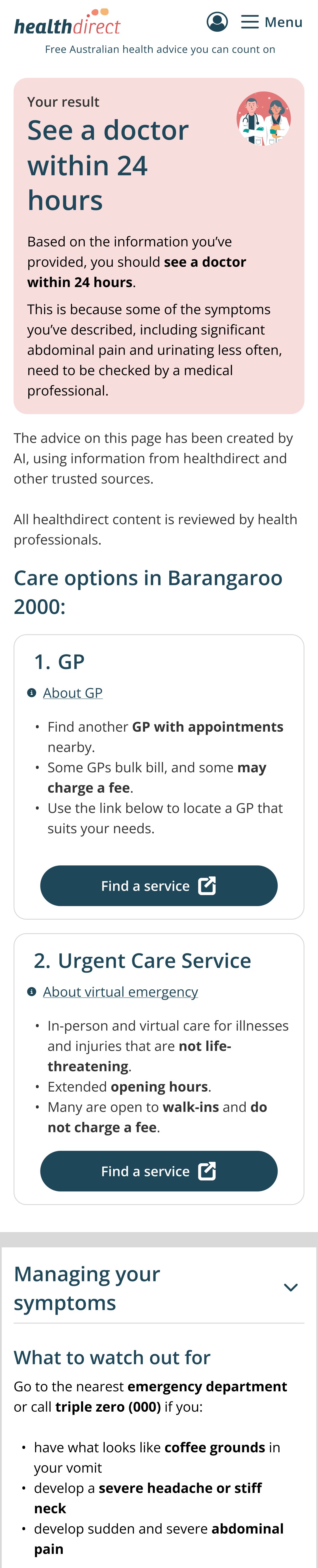

**eFigure 10. Low acuity symptoms, AI-enhanced + detailed information about the use of AI**

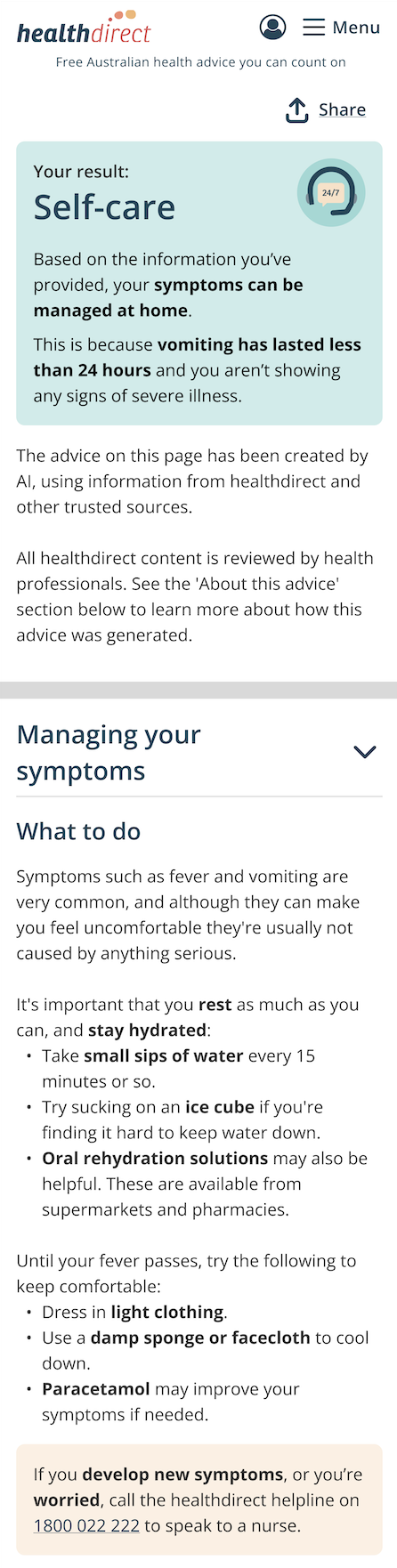

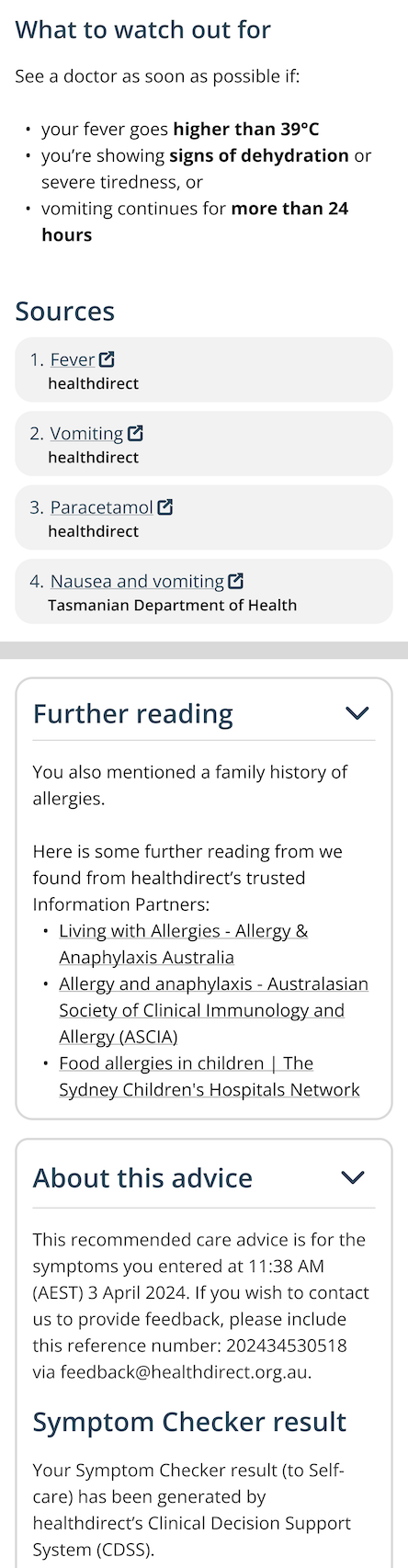

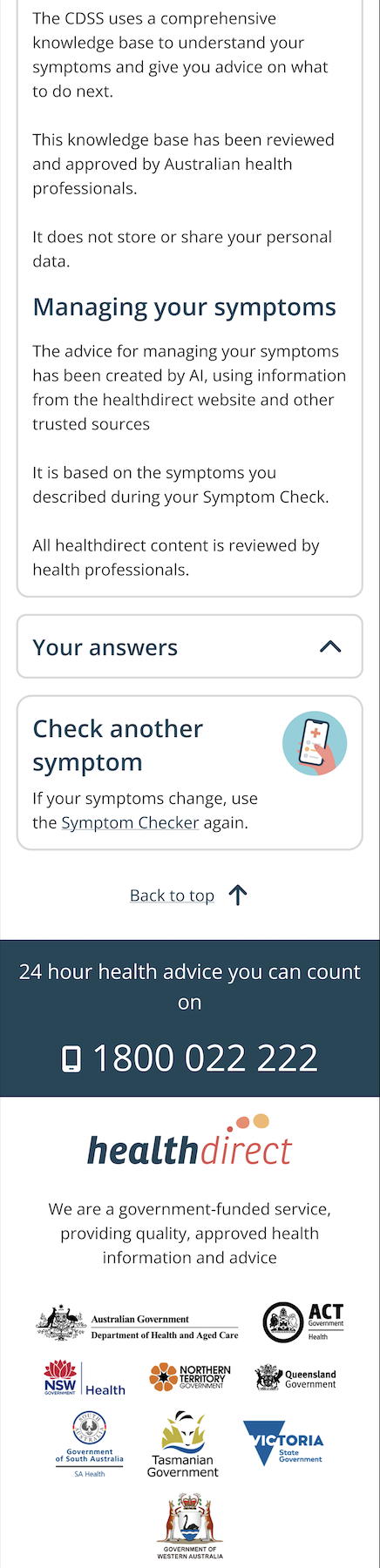

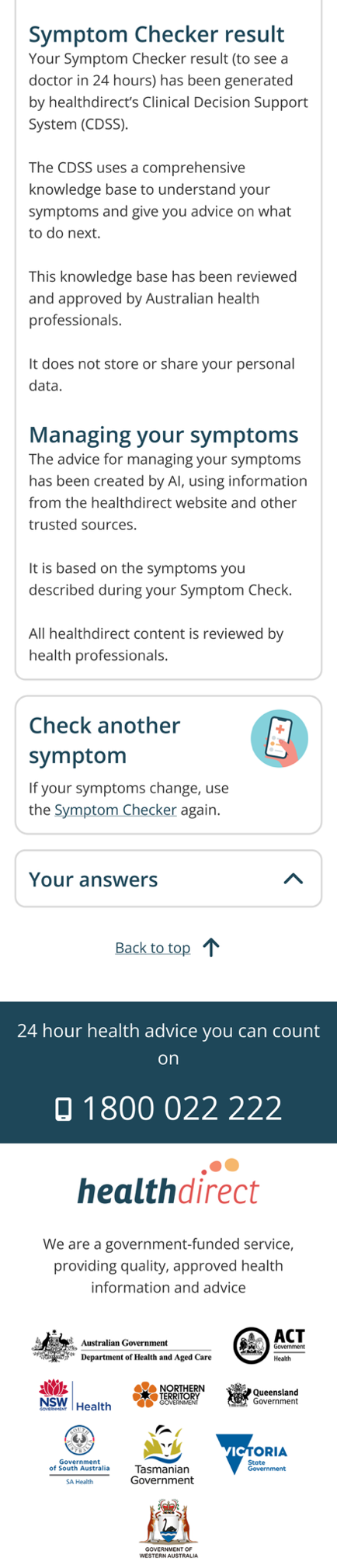

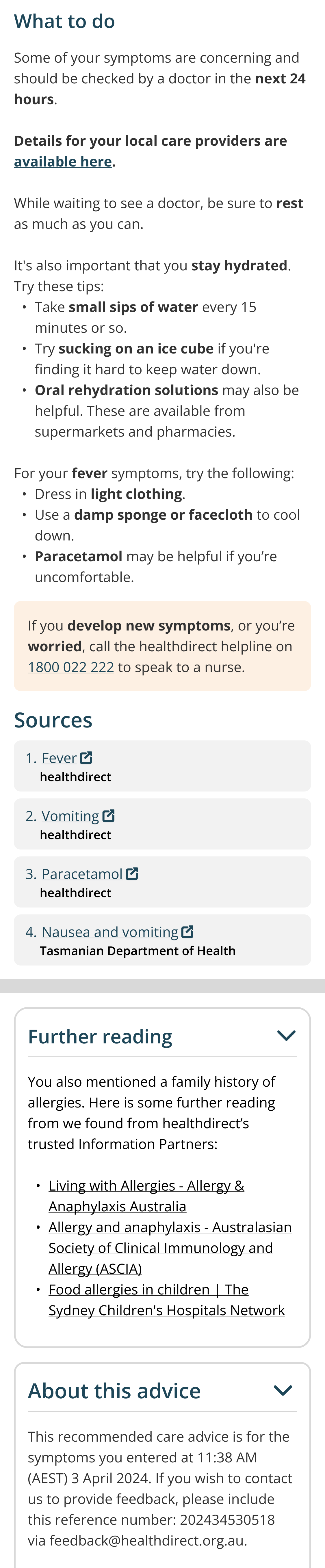

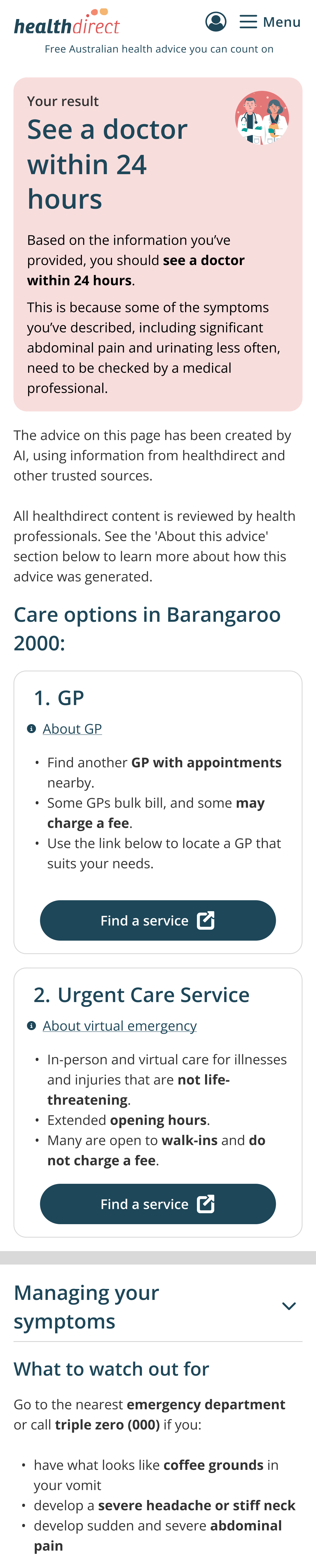

**eFigure 11. Moderate acuity symptoms, AI-enhanced + detailed information about the use of AI**

### Appendix B: Survey items

**eTable 1. Survey items**

| **Demographic survey items** | - Age - Gender - Country of birth - State - Language spoken at home - Aboriginal or Torres Strait Islander - Highest level of education |
| --- | --- |
| **Health Literacy survey item** | - “How often do you need to have someone help you when you read instructions, pamphlets or other written material from your doctor or pharmacy?”   - Always   - Often   - Sometimes   - Occasionally   - Never |
| **Digital health literacy survey item** | - “We would like to ask you for your opinion about your experience using the Internet for health information. For each statement, select which response best reflects your opinion and experience right now.” (From strongly disagree to strongly agree)   - I know **what** health resources are available on the internet.   - I know **where** to find helpful health resources on the internet.   - I know **how** to find helpful health resources on the internet.   - I know **how to use** the internet to answer my questions about health.   - I know how to use the **health information** I find on the internet to help me.   - I have the skills I need to **evaluate** the health resources I find on the internet.   - I can tell **high quality** health resources from **low quality** health resources on the internet.   - I feel **confident** in using information from the internet to make health decisions. |
| **Health survey item** | - “Which, if any, of the following long-standing health conditions do you have (including age-related conditions)?   - Deafness or severe hearing impairment   - Blindness or severe vision impairment   - A long-standing illness (e.g., cancer, HIV, diabetes, chronic heart disease)   - A long-standing physical condition (e.g., arthritis, spinal injury, multiple sclerosis)   - An intellectual disability   - A mental health condition (e.g,, depression)   - A neurological condition (e.g., Alzheimer’s, Parkinson’s)   - None of these |
| **Healthdirect use survey items** | - “Have you heard of Healthdirect before?” - “Which Healthdirect services have you used? (select all that apply)   - Helpline   - Symptom checker   - Web-based information   - Videos   - Service finder   - Mobile app |
| **AI use survey items** | - “How often have you used tools like ChatGPT in the past 6 months?”   - Not at all   - A few times   - Once a month   - Once a week   - Daily - “How often have you used tools like ChatGPT to answer **questions about health** in the past 6 months?”   - Not at all   - A few times   - Once a month   - Once a week   - More than once a week   - Never heard of ChatGPT   - Prefer not to say - “In the last 6 months have you used tools like ChatGPT to . . .”   - Find out what my symptoms mean (or the symptoms of someone I know)   - Find out what to do about a specific health issue that I or someone I know has   - Learn about a specific health condition   - Learn about health lifestyles   - Help create a plan to improve my health (or the health of someone I know)   - Learn more about a medicine, test or treatment (e.g., safety, side effects or interactions)   - Find out if I or someone I know should see a doctor   - Understand medical terms   - Interpret results from blood tests or imaging   - Other (please tell us)   - I have not used tools like ChatGPT to answer health questions |
| **Intention** | - If you were in this situation, would you follow the self-care advice provided   - I would follow all the advice   - I would follow most of the advice   - I would follow some of the advice   - I would follow none of the advice - If you were in this situation, would you make an appointment with your GP in 24 hours?   - Definitely would   - Probably would   - Unsure   - Probably would not   - Definitely would not |
| **Trust** (Mac et al 2024) | - How trustworthy do you find the advice (10-point scale, not trustworthy at all to extremely trustworthy) |
| **Cognitive trust measures** Adapted from Komiak & Benbasat 2006 | - This symptom checker is a real expert in providing relevant health information.” - “This symptom checker has good knowledge.” - “This symptom checker is unbiased.” - “This symptom checker provides me with competent and useful information.” |
| **Emotional trust measures**  Adapted from Komiak & Benbasat 2006 | - “I feel secure about relying on this symptom checker to know what to do next.” - “I feel comfortable about relying on this symptom checker to know what to do next.” - “I feel content about relying on this symptom checker to know what to do next.” |
| **Knowledge** (current symptoms) | - Based on the advice you were given, what can you do to manage your symptoms (select all that apply)   - Drink oral rehydration solution   - Drink small sips of water or suck on an ice cube   - Place a fan in front of you to help cool yourself down   - Consider taking paracetamol if you’re uncomfortable   - Dress in light clothing   - Use a damp sponge or facecloth to help cool yourself down   - Drink hot soup   - Have a cool bath   - None of these   - Don’t know |
| **Knowledge** (low acuity symptoms) | - Based on the advice you were given, when should you seek further medical advice with the GP (select all true statements)   - If you have been experiencing bloody diarrhoea   - If you start dry heaving (dry heaving is when your body tries to vomit but nothing comes out)   - If you notice that your vomit is white and foamy   - If your fever exceeds 39’C   - If your urine turns a darker colour |
| **Knowledge** (moderate acuity symptoms) | - Based on the advice you were given, when should go to the nearest emergency department(select all true statements)   - If your urine starts to turn a darker colour   - If you start dry heaving (dry heaving is when your body tries to vomit but nothing comes out)   - If you notice that your vomit is white and foamy   - If you start to see what resembles coffee grounds in your vomit   - If you develop severe abdominal pain |
| **Acceptability** | - Next time I have symptoms and am unsure what to do, I would use the symptom checker - Would recommend this symptom checker to a friend if they were not sure what to do about symptoms they were experiencing - The symptom checker advice met my expectations   (response options: 5-point Likert scale, strongly disagree to strongly agree). |

### Appendix C: Community involvement

The working group comprised 9 community members who were invited to the project via their involvement in Co-SHeLL. Co-SHeLL is a community panel(1) that supports the work of the Sydney Health Literacy Lab. The nine members had diverse lived experiences across gender, ages, cultural background, with varied health experiences.

Members provided feedback through workshop discussions and email.

**Workshop 1:** Initial activities helped familiarise the working group with the current Healthdirect Symptom Checker tool and described the potential benefits of integrating an AI model with a Retrieval-Augmented-Generation framework into the tool to improve its health advice, thus framing the goals of the project.

This was followed by activities in which working group members discussed their needs, priorities, and expectations for the tool (eBox 1). Discussions also explored how these might change depending on the urgency of the advice.

**Workshop 2:** Insights from these discussions were then incorporated into four prototypes to garner more specific feedback on the design, such as colours, placement, and structure, during a second workshop (eBox 2).

**Post workshop:** Design strategies that the working group and healthdirect staff considered essential were incorporated into the AI-enhanced control. Other design strategies were incorporated into the remaining AI-enhanced intervention groups. Working group insights also identified a clear interest in testing the tool in low and moderate acuity settings and so this was incorporated into the study design.

**eBox 1**: **Summary working group insights (workshop 1)**

| **Potential design strategy** | **Working group insights** |
| --- | --- |
| Empathetic and conversational tone. | - Degree of empathy may need to be tailored to the user (e.g., for neurodivergent users) - Some consumers didn’t perceive the tone as empathetic, but rather neutral - Consider reassurance for highly anxious users |
| Links that are more specific and relate more directly to the content | - Easy to save information and reduces steps required to find the information source - Easy to share with a care provider - Should consider the inclusion of translation services |
| Detailed information about how AI was integrated into the tool | - “Created by AI” may have a negative perception for some people - Include how/when the information was reviewed by health professionals |
| Clear links between the user’s symptoms and acuity level. | - Valued the reasoning/explanation - Keep the reasoning general and AI advice being too specific is perceived as less reliable - Include links to privacy policies |
| **Overall insights**  The working group indicated a strong preference for:   - more video links and visuals - breaking up the advice into different sections - including information about access to user’s local health services - first aid advice in higher acuity settings - more information about where and how user data is stored   The working group described the care advice in the prototype examples as clear and simple, and appropriate for a layperson.   - Need to be specific about whether the advice is a tool for self-assessment (i.e. tailored) or simply a condensed version of information across different websites (not tailored) | |

**eBox 2**: **Summary working group insights (workshop 2)**

| **Discussion stimuli** | **Consumer feedback** |
| --- | --- |
| Provocation 1: symptom checker focused on providing reassurance | - Appreciated the links and disclaimers about where the information has come from - Appreciated the reassurance - Separate the knowledge base sections from the self-care advice |
| Provocation 2: symptom checker focused on the difference between first aid and self-care advice. | - Connect user to local services - “Before your appointment” information should be presented before further information links - Difference between “first aid” and “self-care advice” should be clearer - Blurs the difference between the symptom checker being used as a tool for self-care education vs self-diagnosis |
| Provocation 3: Symptom checker focused on the content structure of the advice | - Monitoring information should be more urgent for “See a GP in 24 hours” acuity - Rationale should be broken up into smaller sections |
| Provocation 4: symptom checker explored different multimedia formats | - Images and videos should be oriented towards action and shouldn’t distract from the important information, but useful for first aid - Videos should be short - Images should be informative rather than promotional - Good for users who struggle with written language - Too much multi-media could be overwhelming for the user |
| **Overall consumer feedback**   - Appreciated the multimedia but should be placed after the most important information, especially for the “See a GP in 24 hours” level of acuity - Multimedia is valuable for technical self-care tips | |

### Appendix D: Analyses

**eTable 1. Participant characteristics at baseline and follow-up (n, %, unless otherwise specified)**

|  | Time point 1 | Time point 2 |
| --- | --- | --- |
|  | **N = 2,062***^1^* | **N = 1,531***^1^* |
| **Age (years)** | 49.00 (35.00, 63.00) | 51.00 (38.00, 65.00) |
| **Gender** |  |  |
| Man | 1,001 (49%) | 723 (47%) |
| Woman | 1,056 (51%) | 805 (53%) |
| Trans and/or gender diverse | 3 (0.1%) | 1 (<0.1%) |
| I use a different term | 1 (<0.1%) | 1 (<0.1%) |
| Prefer not to say | 1 (<0.1%) | 1 (<0.1%) |
| **Born in Australia** | 1,574 (76%) | 1,159 (76%) |
| **What is the main language you speak at home?** |  |  |
| Other | 155 (7.5%) | 119 (7.8%) |
| English | 1,907 (92%) | 1,412 (92%) |
| **Are you of Aboriginal or Torres Strait Islander origin?** |  |  |
| Aboriginal and/or Torres Strait Islander | 66 (3.2%) | 44 (2.9%) |
| No | 1,986 (97%) | 1,479 (97%) |
| Unknown | 10 | 8 |
| **Highest level of education** |  |  |
| No school or other qualifications | 21 (1.0%) | 15 (1.0%) |
| School Certificate or intermediate certificate (or equivalent) | 112 (5.4%) | 82 (5.4%) |
| Higher school certificate or leaving certificate (or equivalent) | 324 (16%) | 213 (14%) |
| Trade Apprenticeship | 93 (4.5%) | 64 (4.2%) |
| Diploma or Certificate | 356 (17%) | 248 (16%) |
| University degree | 1,156 (56%) | 909 (59%) |
| **Health literacy screener** |  |  |
| Adequate health literacy | 1,732 (84%) | 1,306 (85%) |
| Limited health literacy | 330 (16%) | 225 (15%) |
| **eHEALS Score (8-40)** | 31.00 (28.00, 33.00) | 31.00 (28.00, 33.00) |
| **Number of chronic conditions** |  |  |
| 0 | 1,159 (56%) | 859 (56%) |
| 1 | 599 (29%) | 448 (29%) |
| 2 | 211 (10%) | 152 (9.9%) |
| 3+ | 93 (4.5%) | 72 (4.7%) |
| **Healthdirect Use** |  |  |
| Helpline | 243 (12%) | 166 (11%) |
| Symptom Checker | 288 (14%) | 189 (12%) |
| Web-based information | 441 (21%) | 329 (21%) |
| Videos | 87 (4.2%) | 64 (4.2%) |
| Service Finder | 164 (8.0%) | 120 (7.8%) |
| Mobile App | 116 (5.6%) | 83 (5.4%) |
| **How often have you used tools like ChatGPT to answer questions about health in the past 6 months.** |  |  |
| A few times | 435 (21%) | 301 (20%) |
| Once a month | 131 (6.4%) | 91 (5.9%) |
| Once a week | 95 (4.6%) | 67 (4.4%) |
| More than once a week | 79 (3.8%) | 59 (3.9%) |
| Has not used ChatGPT for health | 1,322 (64%) | 1,013 (66%) |

^1^ Median (Q1, Q3)

**eTable 2. Outcomes by format, low acuity symptoms, median (IQR), unless otherwise specified**

|  | **Standard (original tool)** | **AI-enhanced** | **AI-enhanced + Multimedia** | **AI-enhanced + Numbered steps** | **AI-enhanced + Detailed info about AI** | **p-value*^2^*** |
| --- | --- | --- | --- | --- | --- | --- |
|  | **N = 215*^1^*** | **N = 206*^1^*** | **N = 202*^1^*** | **N = 206*^1^*** | **N = 212*^1^*** |  |
| Intent to follow self-care advice (1-4) | 3.00 (1.00), 3.34 | 4.00 (1.00), 3.37 | 3.00 (1.00), 3.32 | 3.00 (1.00), 3.34 | 3.00 (1.00), 3.32 | 0.8 |
| Intent to see a GP in 24 hours (1-5) | 4.00 (1.00), 3.53 | 3.00 (2.00), 3.13 | 3.00 (2.00), 3.37 | 4.00 (2.00), 3.30 | 3.00 (2.00), 3.30 | 0.009 |
| Trust (1-10) mean (SD) | 7.63 (1.42) | 7.61 (1.45) | 7.58 (1.44) | 7.43 (1.73) | 7.49 (1.67) | >0.9 |
| Unknown | 4 | 0 | 2 | 3 | 2 |  |
| Cognitive Trust (4 – 28) mean (SD) | 20.81 (3.82) | 20.67 (3.77) | 20.90 (3.64) | 20.85 (4.44) | 20.69 (4.44) | >0.9 |
| Emotional trust (3-21) mean (SD) | 15.24 (3.59) | 15.43 (3.15) | 15.27 (3.51) | 15.34 (3.94) | 15.29 (3.92) | >0.9 |
| Knowledge for managing current symptoms (%) (immediate) | 0.57 (0.29), 0.58 | 0.86 (0.43), 0.75 | 0.71 (0.43), 0.75 | 0.71 (0.29), 0.73 | 0.86 (0.43), 0.74 | <0.001 |
| Knowledge for managing changing symptoms (%) (immediate) | 0.40 (0.20), 0.53 | 0.80 (0.40), 0.66 | 0.80 (0.40), 0.67 | 0.80 (0.40), 0.65 | 0.80 (0.40), 0.64 | <0.001 |
| Acceptability (1-5) - met expectations | 4.00 (0.00), 3.83 | 4.00 (0.00), 3.92 | 4.00 (0.00), 3.89 | 4.00 (0.00), 3.92 | 4.00 (0.00), 3.89 | 0.7 |
| Acceptability (1-5) - would use the tool in the future. | 4.00 (0.00), 3.84 | 4.00 (0.00), 3.85 | 4.00 (0.00), 3.85 | 4.00 (0.00), 3.86 | 4.00 (1.00), 3.85 | >0.9 |
| Acceptability (1-5) - would recommend the tool | 4.00 (0.00), 3.85 | 4.00 (0.00), 3.86 | 4.00 (1.00), 3.82 | 4.00 (0.00), 3.85 | 4.00 (1.00), 3.78 | 0.9 |

^1^ Median (IQR), Mean

^2^ Kruskal-Wallis rank sum test

**eTable 3. Outcomes by format, moderate acuity symptoms, median (IQR), unless otherwise specified**

|  | **Standard (original tool)** | **AI-enhanced** | **AI-enhanced + Multimedia** | **AI-enhanced + Numbered steps** | **AI-enhanced + Detailed info about AI** | **p-value*^2^*** |
| --- | --- | --- | --- | --- | --- | --- |
|  | **N = 183*^1^*** | **N = 209*^1^*** | **N = 219*^1^*** | **N = 203*^1^*** | **N = 207*^1^*** |  |
| Intent to follow self-care advice (1-4) | 3.00 (1.00) | 3.00 (1.00) | 3.00 (1.00) | 3.00 (1.00) | 3.00 (1.00) | 0.7 |
| Intent to see a GP in 24 hours (1-5) | 4.00 (1.00) | 4.00 (1.00) | 5.00 (1.00) | 4.00 (1.00) | 4.00 (1.00) | 0.08 |
| Trust (1-10) mean (SD) | 7.69 (1.50) | 7.54 (1.71) | 7.72 (1.57) | 7.67 (1.68) | 7.38 (1.64) | 0.2 |
| Unknown | 5 | 9 | 4 | 6 | 3 |  |
| Cognitive Trust (4 – 28) mean (SD) | 21.05 (3.51) | 20.19 (4.26) | 20.81 (3.95) | 20.85 (3.85) | 20.32 (3.96) | 0.4 |
| Emotional trust (3-21) mean (SD) | 15.69 (3.34) | 15.00 (3.81) | 15.16 (3.66) | 15.21 (3.58) | 14.90 (3.50) | 0.3 |
| Knowledge for managing current symptoms (%) | 0.43 (0.43) | 0.71 (0.29) | 0.71 (0.29) | 0.71 (0.29) | 0.71 (0.43) | <0.001 |
| Knowledge for managing changing symptoms (%) | 0.60 (0.40) | 0.80 (0.40) | 0.80 (0.40) | 0.80 (0.40) | 0.80 (0.40) | <0.001 |
| Acceptability (1-5) - met expectations | 4.00 (0.00) | 4.00 (0.00) | 4.00 (0.00) | 4.00 (0.00) | 4.00 (0.50) | 0.4 |
| Acceptability (1-5) - would use the tool in the future. | 4.00 (0.00) | 4.00 (1.00) | 4.00 (1.00) | 4.00 (1.00) | 4.00 (1.00) | 0.4 |
| Acceptability (1-5) - would recommend the tool | 4.00 (1.00) | 4.00 (1.00) | 4.00 (1.00) | 4.00 (1.00) | 4.00 (1.00) | 0.7 |

^1^ Median (IQR), Mean; Mean (SD)

^2^ Kruskal-Wallis rank sum test

**eTable 4. Outcomes at two week follow-up, by format, low acuity symptoms**

|  | **Standard (original tool)** | **AI-enhanced** | **AI-enhanced + Multimedia** | **AI-enhanced + Numbered steps** | **AI-enhanced + Detailed info about AI** |
| --- | --- | --- | --- | --- | --- |
|  | N = 160*^1^* | N = 153*^1^* | N = 158*^1^* | N = 144*^1^* | N = 164*^1^* |
| **Knowledge for managing current symptoms** | |  |  |  |  |
| Immediate (%) | 57.14 (42.86, 71.43) | 85.71 (57.14, 85.71) | 71.43 (57.14, 100.00) | 78.57 (57.14, 92.86) | 71.43 (57.14, 100.00) |
| Follow-up (%) | 57.14 (42.86, 71.43) | 57.14 (42.86, 71.43) | 57.14 (42.86, 71.43) | 57.14 (42.86, 71.43) | 57.14 (42.86, 71.43) |
| *Difference* | *0.00 (-14.29, 14.29)* | *-14.29 (-28.57, 0.00)* | *-14.29 (-28.57, 0.00)* | *-14.29 (-28.57, 0.00)* | *-14.29 (-28.57, 0.00)* |
| **Knowledge for managing changing symptoms** | |  |  |  |  |
| Immediate (%) | 40.00 (40.00, 60.00) | 80.00 (40.00, 80.00) | 80.00 (40.00, 80.00) | 80.00 (40.00, 80.00) | 80.00 (40.00, 80.00) |
| Follow-up (%) | 60.00 (40.00, 80.00) | 60.00 (40.00, 80.00) | 60.00 (40.00, 80.00) | 60.00 (40.00, 80.00) | 60.00 (40.00, 80.00) |
| *Difference* | *0.00 (0.00, 20.00)* | *0.00 (-20.00, 0.00)* | *0.00 (-20.00, 20.00)* | *0.00 (-20.00, 0.00)* | *0.00 (-20.00, 0.00)* |

^1^ Median (Q1, Q3)

**eTable 5. Outcomes at two week follow-up, by format, moderate symptom acuity symptoms**

|  | **Standard (original tool)** | **AI-enhanced** | **AI-enhanced + Multimedia** | **AI-enhanced + Numbered steps** | **AI-enhanced + Detailed info about AI** |
| --- | --- | --- | --- | --- | --- |
|  | N = 122*^1^* | N = 154*^1^* | N = 172*^1^* | N = 158*^1^* | N = 146*^1^* |
| **Knowledge for managing current symptoms** | |  |  |  |  |
| Immediate (%) | 50.00 (28.57, 71.43) | 71.43 (57.14, 85.71) | 71.43 (57.14, 85.71) | 71.43 (57.14, 85.71) | 71.43 (42.86, 85.71) |
| Follow-up (%) | 57.14 (42.86, 71.43) | 57.14 (42.86, 71.43) | 57.14 (42.86, 71.43) | 57.14 (42.86, 71.43) | 57.14 (42.86, 71.43) |
| *Difference* | *0.00 (-14.29, 28.57)* | *-14.29 (-28.57, 14.29)* | *-14.29 (-28.57, 0.00)* | *-14.29 (-28.57, 14.29)* | *-14.29 (-28.57, 14.29)* |
| **Knowledge for managing changing symptoms** | |  |  |  |  |
| Immediate (%) | 60.00 (40.00, 80.00) | 80.00 (60.00, 100.00) | 80.00 (40.00, 80.00) | 80.00 (40.00, 80.00) | 80.00 (40.00, 80.00) |
| Follow-up (%) | 40.00 (40.00, 60.00) | 60.00 (40.00, 80.00) | 40.00 (40.00, 80.00) | 60.00 (40.00, 80.00) | 60.00 (40.00, 80.00) |
| *Difference* | *0.00 (-20.00, 0.00)* | *-20.00 (-40.00, 0.00)* | *-20.00 (-40.00, 0.00)* | *-20.00 (-20.00, 0.00)* | *-10.00 (-20.00, 0.00)* |

^1^ Median (Q1, Q3)

**eTable 6. Outcomes by format, low acuity symptoms, attentive participants**

|  | **Standard (original tool)** | **AI-enhanced** | **AI-enhanced + Multimedia** | **AI-enhanced + Numbered steps** | **AI-enhanced + Detailed info about AI** | **p-value*^2^*** |
| --- | --- | --- | --- | --- | --- | --- |
|  | **N = 208*^1^*** | **N = 194*^1^*** | **N = 193*^1^*** | **N = 197*^1^*** | **N = 201*^1^*** |  |
| Intent to follow self-care advice (1-4) | 3.00 (3.00, 4.00) | 4.00 (3.00, 4.00) | 3.00 (3.00, 4.00) | 3.00 (3.00, 4.00) | 3.00 (3.00, 4.00) | 0.6 |
| Intent to follow advice to see a GP in 24 hours (1-5) | 4.00 (3.00, 4.00) | 3.00 (2.00, 4.00) | 3.00 (2.00, 4.00) | 3.00 (2.00, 4.00) | 3.00 (2.00, 4.00) | 0.008 |
| Trust (1-10) mean (SD) | 7.65 (1.42) | 7.62 (1.44) | 7.57 (1.45) | 7.46 (1.72) | 7.47 (1.67) | 0.9 |
| Unknown | 4 | 0 | 2 | 2 | 2 |  |
| Cognitive Trust (4 – 28) mean (SD) | 20.80 (3.86) | 20.72 (3.82) | 20.87 (3.65) | 20.90 (4.45) | 20.60 (4.43) | 0.9 |
| Emotional trust (3-21) mean (SD) | 15.24 (3.64) | 15.46 (3.19) | 15.28 (3.54) | 15.37 (3.98) | 15.22 (3.92) | 0.9 |
| Knowledge for managing current symptoms (%) | 0.57 (0.43, 0.71) | 0.86 (0.57, 1.00) | 0.71 (0.57, 1.00) | 0.86 (0.57, 1.00) | 0.86 (0.57, 1.00) | <0.001 |
| Knowledge for managing changing symptoms (%) | 0.40 (0.40, 0.60) | 0.80 (0.60, 0.80) | 0.80 (0.40, 0.80) | 0.80 (0.40, 0.80) | 0.80 (0.60, 0.80) | <0.001 |
| Acceptability (1-5) - met expectations | 4.00 (4.00, 4.00) | 4.00 (4.00, 4.00) | 4.00 (4.00, 4.00) | 4.00 (4.00, 4.00) | 4.00 (4.00, 4.00) | 0.5 |
| Acceptability (1-5) - would use the tool in the future. | 4.00 (4.00, 4.00) | 4.00 (4.00, 4.00) | 4.00 (4.00, 4.00) | 4.00 (4.00, 4.00) | 4.00 (3.00, 4.00) | >0.9 |
| Acceptability (1-5) - would recommend the tool | 4.00 (4.00, 4.00) | 4.00 (4.00, 4.00) | 4.00 (3.00, 4.00) | 4.00 (4.00, 4.00) | 4.00 (3.00, 4.00) | 0.8 |

**eTable 7. Outcomes by format, moderate acuity symptoms, attentive participants**

|  | **Standard (original tool)** | **AI-enhanced** | **AI-enhanced + Multimedia** | **AI-enhanced + Numbered steps** | **AI-enhanced + Detailed info about AI** | **p-value*^2^*** |
| --- | --- | --- | --- | --- | --- | --- |
|  | **N = 171*^1^*** | **N = 186*^1^*** | **N = 190*^1^*** | **N = 185*^1^*** | **N = 184*^1^*** |  |
| Intent to follow self-care advice (1-4) | 3.0 (2.0, 4.0) | 3.0 (3.0, 4.0) | 3.0 (3.0, 4.0) | 3.0 (3.0, 4.0) | 3.0 (3.0, 4.0) | 0.5 |
| Intent to follow advice to see a GP in 24 hours (1-5) | 4.0 (4.0, 5.0) | 4.0 (4.0, 5.0) | 5.0 (4.0, 5.0) | 5.0 (4.0, 5.0) | 4.0 (4.0, 5.0) | 0.14 |
| Trust (1-10) mean (SD) | 7.7 (1.5) | 7.5 (1.8) | 7.8 (1.6) | 7.7 (1.6) | 7.4 (1.6) | 0.1 |
| Unknown | 4 | 5 | 4 | 5 | 3 |  |
| Cognitive Trust (4 – 28) mean (SD) | 21.1 (3.5) | 20.2 (4.4) | 20.8 (4.0) | 20.8 (3.7) | 20.5 (3.9) | 0.7 |
| Emotional trust (3-21) mean (SD) | 15.7 (3.4) | 15.0 (3.9) | 15.1 (3.8) | 15.2 (3.6) | 15.0 (3.5) | 0.5 |
| Knowledge for managing current symptoms (%) | 0.4 (0.3, 0.7) | 0.7 (0.6, 0.9) | 0.7 (0.6, 0.9) | 0.7 (0.6, 0.9) | 0.7 (0.4, 0.9) | <0.001 |
| Knowledge for managing changing symptoms (%) | 0.6 (0.4, 0.8) | 0.8 (0.6, 1.0) | 0.8 (0.6, 0.8) | 0.8 (0.6, 0.8) | 0.8 (0.4, 0.8) | <0.001 |
| Acceptability (1-5) - met expectations | 4.0 (4.0, 4.0) | 4.0 (4.0, 4.0) | 4.0 (4.0, 4.0) | 4.0 (4.0, 4.0) | 4.0 (4.0, 4.0) | 0.4 |
| Acceptability (1-5) - would use the tool in the future. | 4.0 (4.0, 4.0) | 4.0 (3.0, 4.0) | 4.0 (3.0, 4.0) | 4.0 (3.0, 4.0) | 4.0 (4.0, 4.0) | 0.3 |
| Acceptability (1-5) - would recommend the tool | 4.0 (4.0, 4.0) | 4.0 (3.0, 4.0) | 4.0 (3.0, 4.0) | 4.0 (3.0, 4.0) | 4.0 (3.0, 4.0) | 0.7 |

^1^ Median (IQR), Mean; Mean (SD)

^2^ Kruskal-Wallis rank sum test

**eTable 8. Sensitivity analysis using multiple imputation**

| **Outcome** | **Multiple Imputation** (n=2546) |  |  | **Complete case**  (n=2062) |  |
| --- | --- | --- | --- | --- | --- |
|  | Median H-statistic  (df=4) | D2 | P-value | H-statistic  (df=4) | P-value |
| Intention to follow self-care advice - **Low Acuity** | 2.351 | F(4, 1620.55)= 0.454 | 0.769 | 1.408 | 0.843 |
| Intention to follow self-care advice - **Moderate Acuity** | 3.106 | F(4, 1498.28)= 0.606 | 0.658 | 2.033 | 0.730 |
| Intention to see a GP in 24 hours - **Low Acuity** | 15.104 | F(4, 889.18)  = 3.039 | 0.017 | 13.417 | 0.009 |
| Intention to see a GP in 24 hours - **Moderate Acuity** | 10.881 | F(4, 898.21)  = 2.068 | 0.083 | 8.349 | 0.080 |

*Notes (eTable 8)*: The data is assumed to be missing at random. Multiple imputation using chained equations with Fully Conditional Specification was performed using the mice package in R version 4.4.3. Ordinal logistic regression was used to impute the missing primary intention variables and 30 datasets were imputed. The imputation model included randomised acuity and presentation, age, sex, country of birth, state of residence, language spoken at home, indigenous status, education level, health literacy screener and number of chronic diseases. Iterations of the mean and standard deviation of the imputed variable were inspected visually and convergence was confirmed. H-statistics were pooled by calculating the D2 statistic and its p values are compared to the complete cases analysis. Median H-statistics from the imputed datasets are also displayed above. The results of the Kruskal Wallis tests performed using multiple imputation overall are comparable to those obtained in the complete case analysis.

To further investigate the “Intention to see a GP” outcome in the low acuity setting, a Dunn test was performed on each of the imputed data sets. The median of the mean ranks in these datasets for this outcome was 694.4, 576.66, 630.7, 659.6 and 623.1 for the control, control AI, detailed AI, multimedia and step-by-step groups respectively. The corresponding mean ranks from the original (smaller) database are 584.5, 485.9, 530.5, 551.0 and 525.9 for each group. The adjusted p values for each pairwise comparison from the imputed datasets was calculated and compared to the original data. Comparable results were found with the only pairwise comparison found to be statistically significant being between the control and control AI groups with a median p value of 0.003 from the imputed datasets, compared to 0.006 in the original analysis.
